# Dismantling cognitive-behavioural therapy for chronic insomnia in adults with or without comorbidities: a systematic review and component network meta-analysis

**DOI:** 10.1101/2023.05.19.23290084

**Authors:** Yuki Furukawa, Masatsugu Sakata, Ryuichiro Yamamoto, Shun Nakajima, Shino Kikuchi, Mari Inoue, Masami Ito, Hiroku Noma, Hikari Nishimura Takashina, Satoshi Funada, Edoardo G. Ostinelli, Toshi A. Furukawa, Orestis Efthimiou, Michael Perlis

## Abstract

**Background:** Chronic insomnia disorder is highly prevalent, disabling and costly. Cognitive-behavioural therapy for insomnia (CBT-I) is recommended as the first-line treatment. CBT-I may contain various educational, cognitive and behavioural strategies delivered in a range of formats, but the effect of each component remains unclear.

**Methods:** We performed a systematic review and component network meta-analysis (cNMA) of CBT-I trials for chronic insomnia. We searched PubMed, CENTRAL, PsycINFO and ICTRP for randomised controlled trials published from database inception to 14^th^ May 2022, comparing any form of CBT-I against each other or a control condition for chronic insomnia disorder in adults (aged ≥18 years). We included insomnia both with and without comorbidities. Concomitant treatments were allowed if they were equally distributed among the arms. Two independent reviewers identified components, extracted data, and assessed trial quality. Primary outcome of interest in this study was treatment efficacy (remission defined as reaching a satisfactory state at endpoint measured by any validated self-reported scale) at post-treatment. (PROSPERO; CRD42022324233)

**Findings:** We identified 226 trials, including 29,982 participants. Mean age was 45·7 years and 71% were women. The results suggests that critical components of CBT-I are cognitive restructuring (incremental odds ratio[iOR] 1·63 [95% confidence interval 1·25 to 2·14]), sleep restriction (iOR 1·44 [1·00 to 2·06]) and stimulus control (iOR 1·44 [1·00 to 2·07]) Sleep hygiene education was not essential (iOR 1·05 [0·79 to 1·38]) and relaxation procedures may be counterproductive (iOR 0·81 [0·64 to 1·03]). Face-to-face, therapist-led program was found to be most beneficial (iOR 1·86 [1·21to 2·85]). The overall risk of bias was low in 8% of the trials, some concerns in 56%, and high in 36%.

**Interpretation:** This cNMA suggests that effective and efficient CBT-I packages can include cognitive restructuring, sleep restriction and stimulus control, but not relaxation.

**Funding:** None.

## INTRODUCTION

Chronic insomnia disorder is characterised by sleep continuity disturbance associated with significant distress or impairments of daytime functioning. It occurs in 4-22% of the population and is highly disabling.^1^ It is associated with both physical and psychiatric comorbidities,^2^ and has a large economic burden.^3^ In 2016, the American College of Physicians recommended that cognitive-behavioural therapy for insomnia (CBT-I) be used as the first-line treatment for chronic insomnia, based on its efficacy and safety profile.^4^ Since then, this recommendation has been supported by several professional societies. ^5, 6^

CBT-I is a structured, non-pharmacological treatment that utilises several educational, cognitive or behavioural strategies for chronic insomnia. Several meta-analyses have shown its effectiveness as a package for chronic insomnia with or without comorbidities.^2, 7, 8^ However, several questions remain open. First, the specific effects of each individual component remain unclear, and it is unknown whether all the components of CBT-I are needed to produce its efficacy. Second, due to its potential for increasing accessibility compared to traditional face-to-face CBT-I, digital CBT-I is becoming more popular, but the best delivery format of CBT-I has not been elucidated.^9^ Few randomised controlled trials (RCTs) have yet examined these component effects and the few available are mostly underpowered. Moreover, combining these studies in meta-analysis is often difficult because they often compared different components to different control conditions. Identifying the effect of each intervention and delivery component is crucial, as it could lead to an intervention that maximises treatment efficacy while minimising the treatment burden and increases its scalability.

Component network meta-analysis (cNMA) is an extension of standard network meta-analysis (NMA)^10^ and can be used to combine all such studies and disentangle the treatment effects of different components included in multicomponent interventions.^11, 12^ In this study, we explored the effect of each treatment component and delivery format of CBT-I for chronic insomnia disorder in adults using cNMA.

## METHODS

The protocol was prospectively registered in PROSPERO (CRD42022324233) and can be found in the appendix (pp 3-12). We report the results following the Preferred Reporting Items for Systematic reviews and Meta-Analyses (PRISMA) guideline extension for NMA.^13^

### Data sources

#### Criteria for considering studies for this review

##### Study design

We included all randomised controlled trials that compared any form of CBT-I against another form of CBT-I or a control condition in the treatment of adults with chronic insomnia. Cluster randomized trials were included in accordance with the Cochrane handbook recommendation^14^ as long as allocation was concealed. If the intra-cluster correlation coefficient was not clearly reported, we assumed it to be 0.05.^15^

##### Participants

We included trials on patients of both genders aged 18 years or older, with chronic insomnia either diagnosed according to formal diagnostic criteria (such as Diagnostic and Statistical Manual of Mental Disorders [DSM]-III-R, DSM-IV, DSM-IV-TR, DSM-5, International Classification of Sleep Disorders [ICSD]-2, ICSD-3, International Classification of Diseases [ICD]-10, ICD-11, or any equivalent criteria^16^), or by elevated scores on self-report measures with satisfactory reliability and validity (such as the Insomnia Severity Index [ISI]). We tested the effect of including studies without a formal diagnosis of insomnia in a sensitivity analysis. We included patients with psychiatric or physical comorbidities ^17^ We tested the effect of including such studies in a sensitivity analysis.

##### Interventions and controls

We regarded CBT-I broadly as a psychotherapy that involved any of the following cognitive or behavioural components: cognitive restructuring, constructive worry, third wave components, sleep restriction, stimulus control, relaxation, and paradoxical intention. Table 1 describes the different components of interest and corresponding prespecified definitions. Co-administration of pharmacological or other psychological therapies was allowed if there were no systematic differences between study arms. CBT-I that clearly included active components focusing on other symptoms such as depression, anxiety or pain was excluded, when such components were not equally distributed among study arms. We included various delivery formats, such as face-to-face contact, individual or group format. In case of self-help program, we included therapeutic guidance and encouragement. We disaggregated these delivery methods as components (Table 1). Control conditions of interest included waiting list, no treatment, attention/psychological placebo control, and treatment as usual. Where multiple arms were reported in a single trial, we included only relevant arms, i.e. arms, that could be described with the components listed in Table 1. We excluded irrelevant arms such as active-drug, pill-placebo, exercise, or bright light therapy with equipment, when such components were not equally distributed among the arms.

**TABLE 1.** List of included components and their definitions

| **Intervention** | **Description** |
| --- | --- |
| **Educational components** |  |
| Sleep hygiene education | General explanation about sleep (e.g., sleep biology, characteristics of healthy sleep, changes in sleep patterns with aging) and general recommendations about lifestyle (e.g., diet, exercise, substance use) and environmental factors (e.g., light, noise, temperature) to improve sleep. This may include some |
|  | elements of other components, e.g. stimulus control, but those should not be the predominant part of the intervention. |
| Sleep diary | Self-monitoring of important daily sleep-related information using sleep diary. |
| **Cognitive components** |  |
| Cognitive restructuring | Skills to identify, challenge and change unhelpful beliefs about sleep that may disturb sleep. Sometimes simply called cognitive therapy. This may include behavioural experiments. |
| Third wave components | Mindfulness and acceptance and commitment therapy. Mindfulness is a form of meditation emphasising a non-judgmental state of heightened or complete awareness of one's thoughts, emotions, or experiences on a moment-to-moment basis. Acceptance and commitment therapy focuses on accepting the feelings and thoughts associated with insomnia through value-based behaviours. |
| Constructive worry | Skills not to worry in bed by writing down the worries and their solutions before going to bed. |
| **Behavioural components** |  |
| Sleep restriction | Skills to improve sleep by limiting time in bed. First, time in bed is restricted to the average sleep duration and then it is increased or decreased depending on sleep efficiency. |
| Stimulus control | Skills to re-associate the bed with sleep. Patients are instructed to; wake up at the same time every morning, refrain from daytime napping, go to bed only when sleepy, get out of bed when unable to sleep, and use the bed/bedroom for sleep and sex only. |
| Relaxation | Structured exercises to reduce somatic tension (e.g., abdominal breathing, progressive muscle relaxation, autogenic training) and cognitive arousal (e.g., guided imagery training). |
| Paradoxical intention | Exercise to remain awake as long as possible after getting into bed. |
| **Others** |  |
| Non-specific treatment effect | Effect of an intervention due to the patients' belief that they are receiving some form of treatment. We classified miscellaneous skills not covered in other sections and not expected to have large effect (e.g., quasi-desensitization) as having non-specific treatment effect. |
| Waiting component | Participants are aware that they can receive an active treatment after a waiting phase. If patients allocated to the waiting list control condition receive some other potentially therapeutic components, we considered both the waiting component and the therapeutic components to be present. |
| Conventional drug treatment | Rated positive when conventional drug treatment is present (drug treatment is |

| Intervention | Description |
| --- | --- |
|  | part of the protocol treatment) or allowed (we will note the percentage of patients on drug). Because this component was always present or absent in each trial, its effect size could not be estimated. |
| **Delivery methods** |  |
| Individual | Individual interaction with therapists, whether it's face-to-face or remote. |
| Group | Interaction with therapists as a member of a group. |
| Face-to-face | Face-to-face interaction with therapists. |
| Therapeutic guidance | Therapeutic guidance in addition to self-help, remote interventions. This may be provided on a scheduled basis or as-needed basis. Technical support only is not included. We coded this component separately from interaction with therapists to see if adding therapeutic guidance to self-help, remote interventions good enough to be effective. |
| Human encouragement | Reminders provided by human beings to proceed with the self-help, remote treatment program via telephone or email. This should not contain any support related to the therapeutic contents. Peer support such as online discussion group was regarded as this component. |
| Automated encouragement | Automated reminders to proceed with the treatment program. This should not contain any support related to the therapeutic contents. |

The components and the names of their combinations were determined based on previous studies ^6, 18^ and content expert consensus among co-authors (MS, RY, SN, TAF, MP).

### Search methods for identification of studies

We conducted a comprehensive literature search in PubMed, the Cochrane Central Register of Controlled Trials (CENTRAL) and PsycINFO on 14^th^ May 2022. We used a combination of index and free terms of psychological treatments and insomnia with filters for randomised clinical trials.^19^ We also searched WHO International Clinical Trials Registry Platform (ICTRP). We imposed no date, language or publication status filters when searching, but included only those that reported sufficient details in English language. We checked the reference lists of review articles for additional potentially eligible records. Appendix (p 7) list the search strings used for each database. We updated our search of trial registries on 6^th^, December 2022.

### Data collection and analysis

#### Selection of studies

Pairs of two reviewers (SF, SK, MS, YF) independently screened titles and abstracts of all potential studies. We then retrieved study reports and pairs of two review authors (RY, SN, MIN, MIT, HN, HNT, SF, SK, MS, YF) independently screened full texts, identified studies for inclusion, and recorded reasons for exclusion of ineligible studies. We resolved any disagreement through discussion or, if required, through consultation with a third reviewer. We identified publications from the same trial so that each trial, rather than each report, was the unit of analysis in our review. We assessed the inter-rater reliability of the full text screening decisions with kappa and percentage agreement.

#### Data items

Pairs of two independent reviewers (RY, SN, MIN, MIT, HN, SK, MS, YF, TAF) extracted data from the included studies, classified all identified treatment arms and their components according to definitions in Table 1 using all available information from publications and inquiries with the original investigators. These pairs assessed validity of the primary outcome of the included trials using the revised risk of bias tool by Cochrane ^20^ in the five following domains: randomisation process, deviations from intended interventions, missing outcome data, measurement of the outcome, and selection of reported results. Any disagreement was resolved through discussion or discussed with a third reviewer if necessary. We measured the inter-rater reliability of the component identification with kappa and percentage agreement, the extracted data concerning the primary outcome with intra-class correlation, and the risk of bias assessment with weighted kappa and percentage agreement.

#### Primary outcome and secondary outcomes

The primary outcome of interest in this study was treatment efficacy (remission defined as reaching a satisfactory state at endpoint measured by any validated self-reported scale) at post-treatment or at its closest time point. If equidistant, we used the longer timeframe. We prioritised inclusion of results from intention-to-treat analyses when possible. When original publications did not report number of remitters, we imputed remission proportions based on continuous outcomes, using a previously validated method.^21^ We assessed the validity of this imputation method by computing the intraclass correlation using studies that report the outcome both in continuous and dichotomous manner. Secondary outcomes included all-cause dropouts (as a proxy measure of treatment acceptability), various subjective sleep parameters including sleep efficiency (%), total sleep time (minutes), sleep onset latency (minutes) and wake after sleep onset (minutes), and long-term remission. We used odds ratio for dichotomous outcomes, standardised mean difference for insomnia severity measures, and mean difference for continuous outcomes expressed in minutes and percent.

#### Statistical analysis

We examined transitivity, which means all the comparisons are on average similar other than the interventions. We created box plots of important trial and patient characteristics to visually examine if potential effect modifiers (publication year, proportion of patients with primary insomnia, age, and baseline severity) were similarly distributed among treatment comparisons. We created a network diagram at the treatment-level to gain a first insight into the network. We checked consistency of the network using global (design-by-treatment) and local (back-calculation) inconsistency tests. ^22, 23^ We then conducted a random-effects, frequentist NMA at the treatment level for the primary outcome.

We visualised NMA results, using psychoeducation as the reference, ranking treatments according to their p-scores.^24^ We also created a league table. We assessed heterogeneity by (i) looking at the standard deviation of random effects (τ^2^) and comparing it against the empirical distributions ^25^; and (ii) creating prediction intervals.^26^ We assessed possible publication bias, reporting bias, and small-study effects using contour-enhanced funnel plots of comparisons with ten or more trials. We investigated the influence of small-study effects in a post-hoc sensitivity analysis excluding arms with less than ten participants. We assessed the certainty of evidence in the treatment-level network estimates of the primary outcome using CINeMA.^27^

We subsequently created a network diagram at the component-level and performed a frequentist random effects cNMA. We used a model that assumes additivity of component effects, i.e. we assumed that the effect of combination therapy is the sum of the effects of its components, and that there are no interactions between them. Given the expected clinical and methodological heterogeneity of treatment effects among the trials, we used the random-effects model. For dropout, we only used studies comparing active treatments because this outcome cannot be meaningfully defined for inactive controls (waiting list, treatment as usual, no treatment). For each component, the cNMA model estimates the incremental odds ratio (iOR), which shows the added benefit or harm of including this component in a combination of other components, and corresponding 95% Confidence Intervals (CIs). To visualise results, we used a method similar to the Kilim plot.^28^ Specifically, we presented each component and corresponding 95% CI using a colouring scheme where green (red) denoted a beneficial (harmful) effect. Shades showed the strength of statistical evidence (p-values of the estimates), with darker shades of green/red indicating stronger statistical evidence in favour/against the corresponding component. White colours indicated absence of evidence. More details are provided in the appendix (p 40). We assessed heterogeneity by calculating the τ^2^ and comparing it against the empirical distributions.^25^

We performed four prespecified sensitivity analyses on the primary outcome by: 1) excluding studies without formal diagnosis of chronic insomnia disorder; 2) excluding studies focusing on patients with comorbidities (both physical and psychological); 3) excluding studies with overall high dropout rate (20% or more); and 4) excluding studies at high overall risk of bias. We conducted the following post-hoc sensitivity analyses, by: 5) focusing on trials that reported ISI; 6) assuming no effect of delivery format components; 7) treating insomnia severity as a continuous outcome; 8) excluding arms with less than 10 participants.

We performed analyses in *R* (version 4.2.3., R foundation, Vienna, Austria)^29^ using the *netmeta* (version 2.8-1) package^30^ to conduct NMA and cNMA, and the *meta* (version 6.2-1) package^31^ to draw contour-enhanced funnel plots.

#### Role of the funding source

There was no funding source for this study.

## RESULTS

### Studies and participants

We identified 7,979 records, assessed 1,709 full-text records for eligibility, and included 226 trials with a total of 29,982 randomised participants (PRISMA flow diagram in appendix p 18). The inter-rater reliability of judgements for full text screening was substantial, with a mean κ of 0·70 and a mean percentage agreement of 85·1%. Appendix (p 19) shows the lists of excluded trials with reasons.

Typical participants were middle-aged women with psychological or physical comorbidities and moderate insomnia symptoms. (mean age 45·7 [SD 16·6] years [221 trials]; 71% female [19,922 of 28,057 reported; 217 trials]; 75% of the trials on insomnia with comorbidities [170 of 226 trials]; mean baseline ISI score 16·7 [SD 4·8; 163 trials]) (Table 2).

**Table 2.** Characteristics of included patients and trials

|  |  |  |
| --- | --- | --- |
| Patient characteristics |  |  |
| Age (year), mean (SD ) | 45.7 (16.6) | 221 trials |
| Gender, N female (%) | 19,922/28,057 (71) | 217 trials |
| Hypnotic use, N (%) | 3,959/19,848 (20) | 159 trials |
| Comorbidities, type | Insomnia without comorbidities | 56 trials |
|  | Psychotic comorbidities | 38 trials |
|  | Physical comorbidities | 71 trials |
|  | Mixed | 61 trials |
| Baseline severity (ISI), mean (SD) | 16.7 (4.8) | 163 trials |
| Diagnosis | Formal operationalised criteria | 138 trials |
|  | Elevated score | 63 trials |
|  | Others | 25 trials |
| Trial characteristics |  |  |
| Region | North America | 117 trials |
|  | Europe | 58 trials |
|  | Asia | 25 trials |
|  | Oceania | 14 trials |
|  | Middle East | 8 trials |
|  | South America | 3 trials |
|  | Africa | 1 trial |
| Number of arms | Two | 194 trials |
|  | Three | 24 trials |
|  | Four | 6 trials |
|  | Five | 1 trial |
|  | Six | 1 trial |
| Randomisation unit | Individual | 224 trials |
|  | Cluster | 2 trials |
| Publication year, mean (SD; range) | 2015 (range; range 1980-2022) | 226 trials |
| Intervention | Total | 494 arms |
| Treatment-level | Cognitive behavioural therapy | 198 arms |
|  | Behavioural therapy | 62 arms |
|  | Cognitive therapy | 12 arms |
|  | No treatment | 3 arms |
|  | Treatment as usual | 8 arms |
|  | Relaxation therapy | 14 arms |
|  | Psychoeducation | 100 arms |
|  | Waiting list | 94 arms |
|  | Attention or psychological placebo | 3 arms |
| Components |  |  |
| Educational components | Sleep hygiene education | 304 arms |
|  | Sleep diary | 409 arms |
| Cognitive components | Cognitive restructuring | 192 arms |
|  | Third wave components | 50 arms |
|  | Constructive worry | 20 arms |
| Behavioural components | Sleep restriction | 231 arms |
|  | Stimulus control | 237 arms |
|  | Paradoxical intention | 19 arms |
|  | Relaxation | 160 arms |
| Others | Non-specific treatment effect | 393 arms |
|  | Waiting component | 98 arms |
| Delivery methods | Individual | 218 arms |
|  | Group | 91 arms |
|  | Face-to-face | 270 arms |
|  | Therapeutic guidance | 38 arms |
|  | Human encouragement | 48 arms |
|  | Automated encouragement | 41 arms |

Operationalised formal diagnostic criteria were used in 138 trials. The 226 included trials had 494 arms (198 cognitive behavioural therapy [CBT] arms; 62 behavioural therapy [BT] arms; 12 cognitive therapy [CT] arms; 3 no treatment [NT] arms; 8 treatment as usual [TAU] arms; 14 relaxation therapy [RT] arms; 100 psychoeducation [PE] arms; 94 waiting list [WL] arms; 3 attention or psychological placebo [PLB] arms). Most trials had two arms (194 two-arm trials; 24 three-arm trials; 6 four-arm trials; 6 four-arm trials; 1 five-arm trials; 1 six-arm trials). We divided five four-arm trials into two two-armed trials in the analysis so that the presence or absence of another concomitant treatment was equally distributed among arms.^32–36^ We also divided one five-arm trial into two trials in the analysis because it reported different outcome measures for subgroups.^37^

Treatment duration ranged from 1 to 16 weeks (median 6 weeks). Hypnotics were used in 3,959 of 19,848 (20%) participants. Study publication year ranged from 1980 to 2022. The inter-rater reliability of judgements for components ranged from moderate to almost perfect, with a mean κ of 0·43 to 0·85 and a mean percentage agreement of 73·1% to 98·8%. (appendix p 21) The inter-rater reliability of extracted data for the primary outcome was almost perfect, with an intraclass correlation (ICC) of 0·98. Validity of the remission imputation method was confirmed, with an ICC of 0·96 (appendix p 22).

The overall risk of bias according to the Cochrane’s revised risk of bias tool was low in 17 trials (8%, 17/226), some concerns in 127 (56%, 127/226) and high in 82 (36%, 82/226). The inter-rater reliability for the overall risk of bias was fair, with a mean weighted κ of 0·32 and a mean percentage agreement of 52·9% (the mean weighted κ for each domain ranged from 0·13 to 0·55; the mean percentage agreement from 53·4% to 77·8%). (appendix p 23)

Assessment of transitivity (appendix p 27) showed that potential effect modifiers were evenly distributed across comparisons, except for the proportion of insomnia without comorbidities. We explored its impact in a prespecified sensitivity analysis (details below).

### Treatment-level network meta-analysis

The network for the primary outcome on the treatment-level was well-connected. (Figure 1) The global test did not show evidence of inconsistency (p=0·20). The back-calculation method identified only two comparisons out of 20 that suggested inconsistency, a proportion that would be empirically expected.^38^ We report our evaluation of inconsistency for the primary outcome in the appendix (p 28)

**Figure 1.**
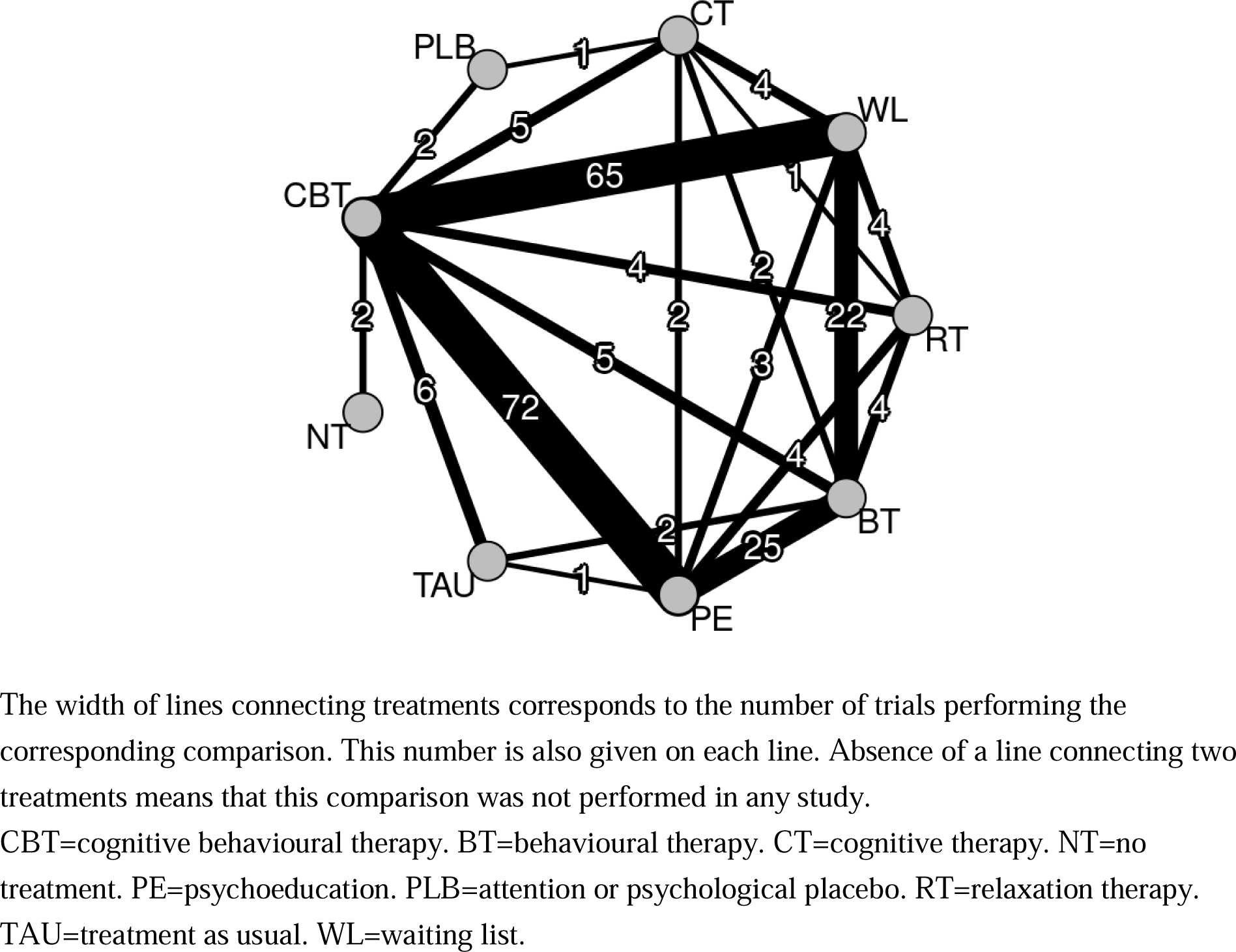
Network diagram

Figure 2 shows the results of the treatment-level NMA, with treatments ranked according to their p-scores. CBT was the most efficacious treatments, followed by BT and CT, suggesting the effectiveness of both cognitive and behavioural skills. In the appendix (p 30) we provide the league table.

**Figure 2.**
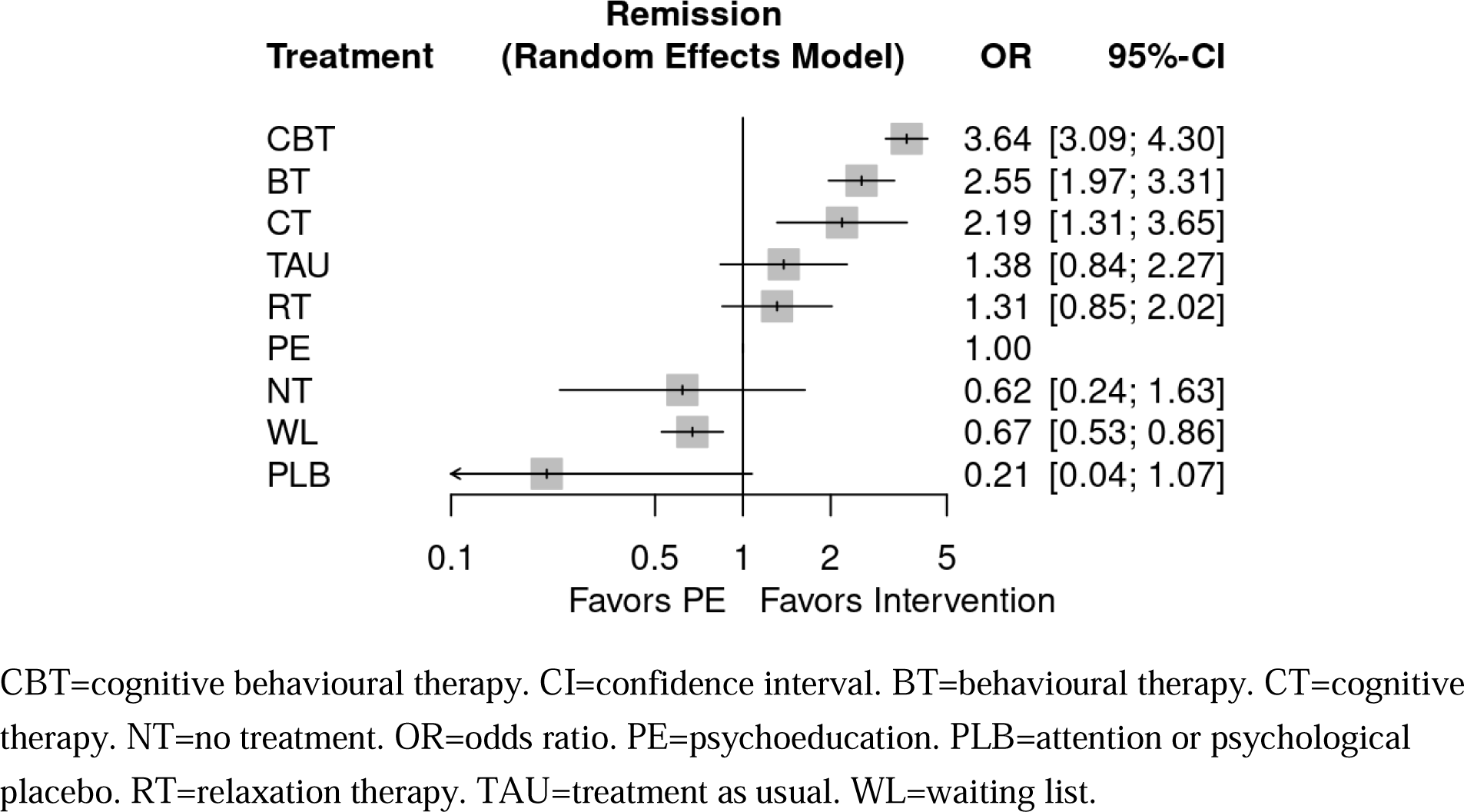
Relative effects of treatments versus psychoeducation as estimated from network meta-analysis

The heterogeneity of the primary outcome was within the empirically expected range (τ^2^ =0.21).^25^ The prediction interval did not change the overall interpretation of the results (appendix p 31) The contour-enhanced funnel plots suggested no publication or reporting biases. However, we found some evidence of possible small study effects, i.e. smaller studies sometimes showed consistently different results than bigger ones (appendix p 33). We explored the impact of this in a post-hoc sensitivity analysis (details below). Moderate certainty of evidence suggested the superiority of CBT, BT and CT over PE, and low certainty of evidence CBT over BT and CT. Other combinations were of very low to moderate certainty. Appendix (p 35) provides full information on CINeMA.

### Component network meta-analysis

We then conducted the component network meta-analysis to assess the effectiveness of each component. The component-level network was well-connected. (appendix p 39). Figure 3 shows primary and secondary outcomes for all components. The heterogeneity was again within the empirically expected range (τ^2^=0.32).^25^

**Figure 3.**
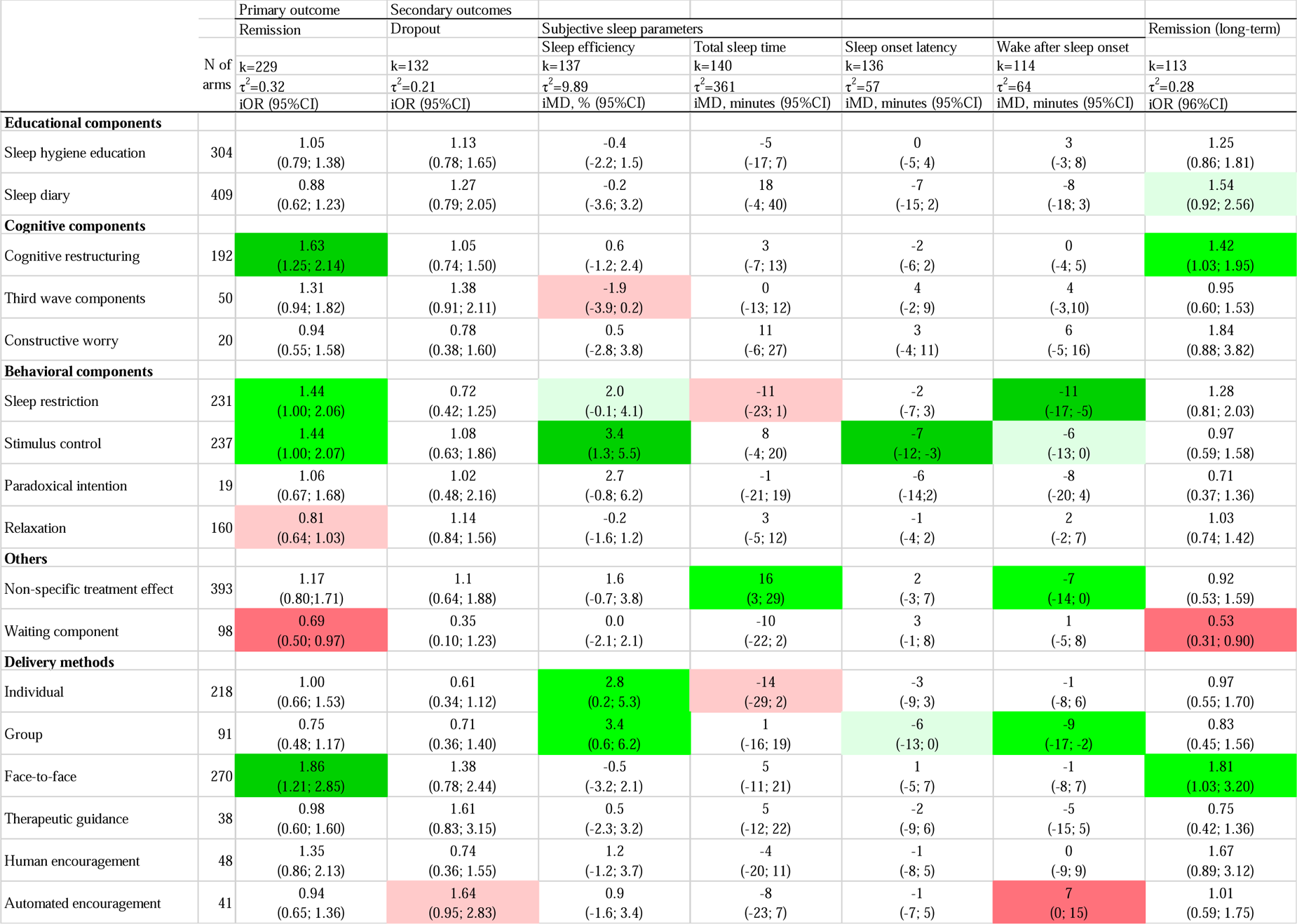
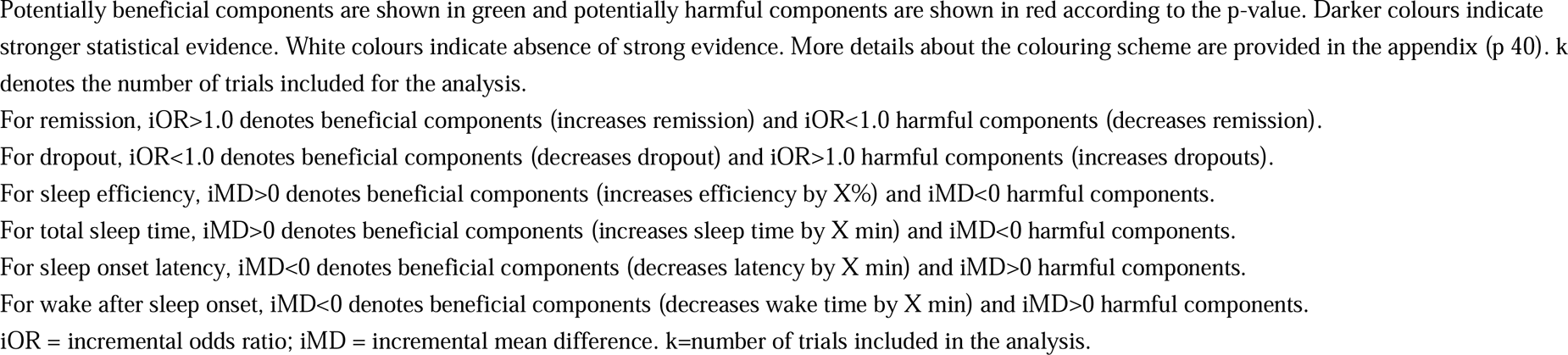
Primary and secondary outcomes of component network meta-analysis

The cNMA revealed that cognitive restructuring, sleep restriction and stimulus control may have beneficial effects, but relaxation might be detrimental for the primary outcome. Face-to-face contact with the therapists may also be helpful. The waiting component decreased the remission rate, and thus may inflate the efficacy estimate of the active arm when used as the control.

There were no components that importantly increased or decreased acceptability of the treatment. In terms of subjective sleep parameters, sleep restriction improved wake after sleep onset, while stimulus control improved sleep efficiency and sleep onset latency. We did not find evidence suggesting an effect of cognitive restructuring and face-to-face delivery on sleep parameters. These two components remained efficacious in terms of long-term remission. (Figure 3)

Results of pre-specified and post-hoc sensitivity analyses largely confirmed our primary analyses. (appendix p 41)

## DISCUSSION

We conducted the first systematic review and cNMA of CBT for chronic insomnia in adults. At the treatment-level, we found that CBT was the most efficacious treatment package followed by BT and CT. At the component-level, we found that cognitive restructuring, sleep restriction and stimulus control may be beneficial, while relaxation might be harmful. With regard to the delivery format, individual or group face-to-face sessions may be beneficial.

Our results also showed that these components have target-specific effects on the secondary outcomes. Cognitive restructuring improved quality of sleep without significantly changing sleep parameters and its effect was long-lasting. Sleep restriction therapy improved wake after sleep onset, and stimulus control therapy improved sleep onset latency. This supports the notion that BT and CT have different mechanisms of actions,^39^ and encourage the use of cognitive approach for insomnia,^40^ despite the recent trend of behavioural centred treatment.^41^ Treatment manuals state that there is no evidence to indicate that these components are differentially effective for specific types of insomnia,^42, 43^ and the meta-analyses by AASM in 2021 were also inconclusive about the specific sleep parameters of singe-component strategies. While the current study only used aggregate data and could not examine individual factors affecting treatment efficacies, such differential effects as we found in this study may lead to more personalised treatments for insomnia by matching individuals’ insomnia characteristics with appropriate techniques.

Sleep hygiene education and sleep diary, which are included in most of the CBT-I packages, may not be beneficial or harmful on their own. On the other hand, our cNMA found relaxation component, which is also often part of the CBT-I packages, to be potentially harmful. The effectiveness of relaxation therapy may not be because of the effectiveness of relaxation techniques, but because of the non-specific treatment effect and the face-to-face component it often includes. It should be noted that, in the treatment of insomnia, relaxation might lead to lying longer while awake, which might work in the opposite direction of sleep restriction or stimulus control. As a matter of fact, the cNMA of CBT for panic disorder found muscle relaxation as potentially harmful,^44^ and the cNMA of internet CBT for depression also found relaxation to be potentially detrimental.^18^ Furukawa and colleagues^18^ argued that relaxation might have worked in the opposite direction of exposure technique, which might be the fundamental treatment component for panic disorder. In the treatment of depression, relaxation might have worked in the opposite direction of behavioural activation, which might be the principal therapeutic mechanism in depression.^18^

Our results were in agreement with previous systematic reviews and meta-analyses in that individual or group face-to-face delivery formats are the most efficacious, and guided or unguided self-help can be a viable alternative.^45, 46^

Our study has several limitations. First, our analysis considered components either present or absent based on the descriptions provided in each study. In reality, components may have varied between programmes in terms of their contents and the extent to which they were implemented. Second, we assumed additivity of the components effect and no interactions between components. Third, the high dropout rate of 26% (7,581 of 29,166) may have influenced the results. We made our analysis conservative by treating those dropped out from assessment as non-remitters. We conducted a sensitivity analysis excluding trials with overall high dropout rate (20% or more) and the results were consistent with the primary analysis.

By contrast, the strengths of this study may be as follows. First, we performed a comprehensive and up-to-date systematic review and included 226 trials, more than twice the number of trials included in the meta-analyses by AASM.^8^ To our knowledge, this study is by far the largest meta-analysis ever done in the field of CBT-I, and the largest cNMA in psychotherapies.^18, 44, 47^ Second, we used the cutting-edge method to assess the specific efficacies of various skills and delivery formats of CBT-I. Identification of the components is a key in conducting the cNMA. We a priori made a table of the definitions of components (Table 1), and achieved moderate to almost perfect inter-rater agreement (appendix p 21). Although the statistical indices of heterogeneity of our cNMA were not smaller than those of our NMA, they were both within the empirically expected range.^25^

In conclusion, most efficacious and efficient CBT-I package may include cognitive restructuring, sleep restriction and stimulus control in the face-to-face individual or group format. They all contribute to the perceived quality of sleep: cognitive restructuring and face-to-face contact mainly affect the quality of sleep both at short-term and long-term, while sleep restriction and stimulus control, but not cognitive restructuring and face-to-face contact, affect sleep parameters. Sleep hygiene education may not be essential, and relaxation may be harmful. To increase scalability, self-help program with human encouragement warrant further trials. We hope that this study will help clinicians provide patients with effective treatments with less burden. Policy makers and guideline developers can utilise the results of this study to facilitate the dissemination of CBT-I efficiently.

## Data Availability

All data produced in the present study are available upon reasonable request to the authors

## Contributors

YF and MS conceived the study. YF, MS, EGO, TAF, MP and OE designed the study. YF, MS, SK, and SF independently screened titles and abstracts of all potential studies, YF, MS, RY, SN, SK, MIN, MIT, HN, HNT, and SF screened full texts. YF, MS, RY, SN, MIN, MIT, HN, SK, and TAF independently extracted data. YF and OE analysed the data. YF, MS, TAF, OE and MP interpreted the results. YF wrote the first draft of the manuscript. All authors had access to all the data and provided critical input and revisions to the draft manuscripts and approved the full manuscript.

## Declaration of interests

YF has received consultancy fee from Panasonic outside the submitted work.

MS reports personal fees from SONY outside the submitted work.

SF received a research grant from JSPS KAKENHI (Grant Number JP 20K18964) and the Pfizer Health Research Foundation outside this project.

EGO has received research and consultancy fees from Angelini Pharma.

TAF reports personal fees from Boehringer-Ingelheim, DT Axis, Kyoto University Original, Shionogi and SONY, and a grant from Shionogi, outside the submitted work; In addition, TAF has patents 2020-548587 and 2022-082495 pending, and intellectual properties for Kokoro-app licensed to Mitsubishi-Tanabe.

OE was supported by the Swiss National Science Foundation (Ambizione grant number 180083) MP wrote treatment manuals and books for CBT-I, teaches CBT-I, and is a founder of Hypknowledge LLC.

RY, SN, SK, MIN, MIT, HN, HNT, OE report no competing interest.

## Data sharing

Data and code used for analyses are available from the corresponding author on reasonable request.

## Acknowledgement

The views expressed are those of the authors and not necessarily those of affiliated organizations. We thank Ms Hanae Daimon for helping us retrieve full texts

## Registration

This protocol is prospectively registered in PROSPERO (CRD42022324233).

This research was prospectively registered (#2022033NIe), Ethical Committee, Faculty of Medicine, The University of Tokyo.

## Notes

### Clinical Protocols

https://www.crd.york.ac.uk/prospero/display_record.php?RecordID=324233

### Funding Statement

This study did not receive any funding

