## Appendix for "Dismantling cognitive-behavioural therapy for chronic insomnia in adults with or without comorbidities: a systematic review and component network meta-analysis"

### **1. Protocol**

We prospectively registered the protocol in PROSPERO ([CRD42022324233](https://www.crd.york.ac.uk/prospero/display_record.php?RecordID=324233)) on 10 May 2022. You can find the full text protocol on the preprint server (Furukawa Y, et al. medRxiv. 2022. doi: <https://doi.org/10.1101/2022.06.02.22275890> ) Here we present the protocol as of 25 May, 2022 and the changes.

#### 1.1. Protocol as of 25 May, 2022

First draft: 4 March, 2022

Last update before PROSPERO registration: 23 April, 2022

Last updated:　25 May, 2022

**Dismantling cognitive-behavioral therapy for chronic insomnia: a protocol for a systematic review and component network meta-analysis**

**REVIEW QUESTION**

What is the effect of each component of cognitive behavioral therapy for chronic insomnia?

**BACKGROUND**

Insomnia is highly prevalent and disabling.(Roth et al., 2011) Clinical practice guidelines recommend cognitive-behavioral therapy for insomnia (CBTI) as the first-line treatment.(Edinger et al., 2021; Qaseem et al., 2016) CBTI includes various combinations of many different components and its clinical benefits have been shown as a package,(Straten et al., 2018) whereas the effect of each component remains unclear. Finding the effect of each component will lead to an intervention that may maximize treatment benefit while reducing the number of components, thereby reducing the treatment burden, lowering the cost of CBTI and making it available to more people. Component network meta-analysis (CNMA) is an extension of standard network meta-analysis that can be used to disentangle the treatment effects of different components included in multicomponent interventions. (Rücker et al., 2020b) In this study, we will explore the effect of each component of CBTI with the use of CNMA.

**METHODS**

We will follow the Preferred Reporting Items for Systematic reviews and Meta-Analyses (PRISMA) guideline extension for NMA. (Hutton et al., 2015) The protocol is prospectively registered in PROSPERO (CRD42022324233).

**Data sources**

**Criteria for considering studies for this review**

***Study design***

We will include all randomized controlled trials that compared any form of CBTI against another form of CBTI or a control condition in the treatment of adults with chronic insomnia. Cluster randomized trials will be included in meta-analyses in accordance with the Cochrane handbook recommendation. (Higgins et al., 2022) If the intra-cluster correlation coefficient is not clearly reported, we assume it to be 0.05.(Lin et al., 2018)

***Participants***

We will include studies on patients of both genders aged 18 years or older with insomnia either diagnosed according to formal diagnostic criteria (such as the Diagnostic and Statistical Manual of Mental Disorders, the International Classification of Diseases or the International Classification of Sleep Disorders) or judged so by clinical expertise (e.g. presence of significant symptoms). The effect of including studies without a formal diagnosis of insomnia will be tested in a sensitivity analysis. We will include patients with psychiatric or physical comorbidities. (Sateia, 2014) The effect of including such studies will be examined in a sensitivity analysis.

***Interventions and controls***

We regard CBTI broadly as a psychotherapy involving any one of the following cognitive or behavioral components. Table 1 describes the different components of interest and their definitions. Pharmacological co-administration will be allowed as long as it is stated that there were no systematic differences in drug administration between study arms. CBTI that clearly includes active components focusing on other symptoms, such as depression, anxiety or pain, will be excluded.

The control conditions of interest will include waiting list, no treatment, attention/psychological placebo control and treatment as usual. In this CNMA study, treatment as usual must include pharmacotherapy; watchful waiting will be classified as attention/psychological placebo even when it is termed ‘treatment as usual’ in some papers. When no treatment, attention/psychological placebo or treatment as usual are used while on the waiting list, such controls will be regarded as waiting list in the NMA but will be decomposed as appropriate in the CNMA. We will include interventions of any duration.

Where multiple arms are reported in a single trial, we will include only the relevant arms that can be described with the components listed in Table 1. We will lump the arms when they share the same components (e.g. same components delivered in 30 min per session against 60 min per session). We will exclude irrelevant arms such as active-drug, pill-placebo, exercise or bright light therapy with equipment.

Possible components and their combinations are shown in Table 1 and Table 2. The components and nodes were determined based on previous studies (Edinger et al., 2021; Furukawa et al., 2021) and content expert consensus among co-authors (MS, TAF, MP).

**TABLE 1 List of included components and their definitions**

| Intervention | Description |
| --- | --- |
| Educational components (EDU) | |
| Sleep hygiene education (se) | General explanation about sleep (eg, sleep biology, characteristics of healthy sleep, changes in sleep patterns with aging, stress biology, and its impact on sleep) and general recommendations about lifestyle (eg, diet, exercise, substance use) and environmental factors (eg, light, noise, temperature) to improve sleep. This may include some elements of other components but those should not be the predominant part of the intervention. |
| Sleep diary (sd) | Self-monitoring of important daily sleep-related information using diary. |
| Cognitive components (COG) | |
| Cognitive restructuring (cr) | Skills to identify, challenge and change unhelpful beliefs about sleep that may disturb sleep. This may include behavioral experiments. |
| Mindfulness (mi) | A form of meditation emphasizing a nonjudgmental state of heightened or complete awareness of one’s thoughts, emotions, or experiences on a moment-to-moment basis. |
| Constructive worry (cw) | Skills not to worry in bed by writing down the worries and their solutions before going to bed. |
| Imagery rehearsal (ir) | Skills to identify, challenge and change nightmares that disturb sleep. |
| Behavioral components (BEH) | |
| Sleep restriction (sr) | Skills to improve sleep by limiting time in bed. First, time in bed is restricted to the average sleep duration and then it is increased or decreased depending on sleep efficiency. |
| Stimulus control (sc) | Skills to re-associate the bed with sleep. Patients are instructed to; wake up at the same time every morning, refrain from daytime napping, go to bed only when sleepy, get out of bed when unable to sleep, and use the bed/bedroom for sleep and sex only. |
| Relaxation (re) | Structured exercises designed to reduce somatic tension (eg, abdominal breathing, progressive muscle relaxation, autogenic training) and cognitive arousal (eg, guided imagery training). |
| Paradoxical intention (pi) | Exercise to remain awake as long as possible after getting into bed. |
| Others | |
| Waiting component (w) | Participants are aware that they can receive an active treatment after a waiting phase. If patients allocated to the waiting list control condition receive some other potentially therapeutic components, we will consider both the waiting component and the therapeutic components to be present. |
| Conventional drug treatment (dt) | Rated positive when conventional drug treatment is present (drug treatment is part of the protocol treatment) or allowed (we will note the percentage of patients on drug). |
| Non-specific treatment effect (ns) | Effect of an intervention due to the patients’ belief that they are receiving some form of treatment. Miscellaneous skills not covered in other sections and not expected to have large effect (i.e. quasi-desensitization) are classified as having non-specific treatment effect. |
| Self-help, unguided, remote (NA) | Default delivery format is self-help, unguided and remote, as it requires the least human resource. |
| Human encouragement (he) | Reminders provided by human beings to proceed with the self-help, remote treatment program via telephone or email. This should not contain any support related to the therapeutic contents. Peer support such as discussion group will be regarded as this component. We will code this component separately from interaction with therapists (in, gp, ff) to see if adding human encouragement to self-help, remote interventions good enough to be effective. |
| Therapeutic guidance (tg) | Therapeutic guidance in addition to self-help, remote interventions. This may be provided on a scheduled basis or as-needed basis. Technical support only is not included. We will code this component separately from interaction with therapists (in, gp, ff) to see if adding therapeutic guidance to self-help, remote interventions good enough to be effective. |
| Individual (in) | Individual interaction with therapists. The combination (in + ff) means individual, face-to-face sessions are held. The combination (in – ff) individual remote sessions, such as via telephone or videoconference. |
| Group (gp) | Interaction with therapists as a member of a group. |
| Face-to-face (ff) | Face-to-face interaction with therapists. |
| Automatic encouragement (ae) | Automated reminders to proceed with the treatment program. This can be added both to the self-help interventions or the interventions including interactions with therapists. This should not contain any support related to the therapeutic contents. |

**TABLE 2 Conceptualization of cognitive behavioral therapy for insomnia or control conditions from the component perspective**

|  | **Possible combinations of components** |
| --- | --- |
| Cognitive behavioral therapy | + ns ± EDU + COG + BEH ± dt ± he ± tg ± in ± gp ± ff ± ae |
| Cognitive therapy | + ns ± EDU + COG ± dt ± he ± tg ± in ± gp ± ff ± ae |
| Behavioral therapy | + ns ± EDU + BEH ± dt ± he ± tg ± in ± gp ± ff ± ae |
| Psychoeducation | + ns + EDU ± dt ± he ± tg ± in ± gp ± ff ± ae |
| Waiting list | + w ± ns ± EDU ± dt ± he ± tg ± in ± gp ± ff ± ae |
| Treatment as usual | + ns + dt + in + ff |
| Attention or psychological placebo | + ns ± dt ± he ± tg ± in ± gp ± ff ± ae |
| No treatment | ± ff ± in |

Components marked with a “+” are required. Components marked with “±” are optional. Components not mentioned cannot be included. Capitalized components (EDU, COG, BEH) mean at least one of the components in that group is required. ae=automatic encouragement. BEH=behavioral components. COG=cognitive components. dt=conventional drug treatment. EDU=educational components. ff=face-to-face. gp=group. he=human encouragement. in=individual. ns=non-specific treatment effect. tg=therapeutic guidance. w=waiting component.

**Search methods for identification of studies**

We will carry out a comprehensive literature search in PubMed, CENTRAL and PsycINFO. We will use a combination of index and free terms of psychological treatments and insomnia with filters (Eady et al., 2008) for randomized clinical trials. We will also search WHO International Clinical Trials Registry Platform. We will impose no date, language or publication status restriction. We will check the reference lists of review articles for additional potentially eligible records.

**TABLE 3 Search strings for PubMed, Cochrane Central Register of Controlled Trials and PsycINFO**

| Database | Search strings |
| --- | --- |
| PubMed | (Psychotherapy [MH] OR psychotherap*[All Fields] OR "cognitive behavioural therapy"[All Fields] OR "cognitive behavioral therapy"[All Fields] OR "CBT"[All Fields] OR "CBTI"[All Fields] OR "CBT-I"[All Fields] OR "cognitive therapy"[All Fields] OR "behavioural therapy"[All Fields] OR "behavioral therapy"[All Fields] OR "sleep hygiene"[All Fields] OR "sleep education"[All Fields] OR "sleep diary"[All Fields] OR "cognitive restructuring"[All Fields] OR "mindfulness"[All Fields] OR "constructive worry"[All Fields] OR "imagery rehearsal"[All Fields] OR "relaxation"[All Fields] OR "sleep restriction"[All Fields] OR "stimulus control"[All Fields] OR "paradoxical intention"[All Fields])  AND  ("sleep wake disorders"[MeSH Terms] OR insomnia[All Fields])  AND  (randomized controlled trial[pt] OR controlled clinical trial[pt] OR randomized[tiab] OR placebo[tiab] OR clinical trials as topic[mesh:noexp] OR randomly[tiab] OR trial[ti] NOT (animals[mh] NOT humans [mh])) |
| Cochrane Central Register of Controlled Trials | ([mh Psychotherapy] OR psychotherap* OR "cognitive behavioural therapy" OR "cognitive behavioral therapy" OR "CBT" OR "CBTI" OR "CBT-I" OR "cognitive therapy" OR "behavioural therapy" OR "behavioral therapy" OR "sleep hygiene" OR "sleep education" OR "sleep diary" OR "cognitive restructuring" OR mindfulness OR "constructive worry" OR "imagery rehearsal" OR relaxation OR "sleep restriction" OR "stimulus control" OR "paradoxical intention")  AND  ([mh "sleep wake disorders"] OR insomnia) |
| PsycINFO | (exp Psychotherapy/ OR psychotherap*.af. OR "cognitive behavioural therapy".af. OR "cognitive behavioral therapy".af. OR "CBT".af. OR "CBTI".af. OR "CBT-I".af. OR "cognitive therapy".af. OR "behavioural therapy".af. OR "behavioral therapy".af. OR "sleep hygiene".af. OR "sleep education".af. OR "sleep diary".af. OR "cognitive restructuring".af. OR mindfulness.af. OR "constructive worry".af. OR "imagery rehearsal".af. OR relaxation.af. OR "sleep restriction".af. OR "stimulus control".af. OR "paradoxical intention".af.)  AND  (exp "sleep wake disorders"/ OR insomnia.af.)  AND  ("double-blind" OR "random* assigned" OR control) |
| WHO International Clinical Trials Registry Platform | (psychotherap* OR "cognitive behavioural therapy" OR "cognitive behavioral therapy" OR "CBT" OR "CBTI" OR "CBT-I" OR "cognitive therapy" OR "behavioural therapy" OR "behavioral therapy" OR "sleep hygiene" OR "sleep education" OR "sleep diary" OR "cognitive restructuring" OR mindfulness OR "constructive worry" OR "imagery rehearsal" OR relaxation OR "sleep restriction" OR "stimulus control" OR "paradoxical intention")  AND  ("sleep wake disorders" OR insomnia) |

**Data collection and analysis**

**Selection of studies**

Two review authors will independently screen titles and abstracts of all the potential studies we identify as a result of the search and code them as ‘retrieve’ or ‘do not retrieve’. We will retrieve the full text study reports/publications and two review authors will independently screen the full text and identify studies for inclusion and identify and record reasons for exclusion of the ineligible studies. We will resolve any disagreement through discussion or, if required, we will consult a third reviewer. We will identify publications from the same study so that each study rather than each report is the unit of analysis in the review. We will record the selection process in sufficient detail to complete a PRISMA flow diagram.

**Data items**

Two review authors will extract independently data from the included studies. Any disagreement will be resolved through discussion, or discussed with a third person if necessary. We will abstract the following information.

***1. Characteristics of the studies***

Name of the study, year of publication, country, study site (single or multi-center), study design (individually-randomized or cluster-randomized), population characteristics (mean age, number of women, definition of insomnia, number of patients with primary insomnia), intervention (components, delivery format, qualification of therapists [if applicable], duration), outcomes (scale used for the primary outcome)

***2. Identification of components***

Two independent reviewers will determine the classification of all identified arms and their components according to the definitions in Table 1, based on all available information including the publications, trial registry and inquiry with the original investigators if needed. Any disagreement will be solved by the two reviewers and, where necessary, in consultation with a third member of the review team. We will report the inter-rater agreement in terms of percentage agreement and kappa.

***3. Risk of bias***

We will use Cochrane Risk of Bias 2.0 tool (RoB2) (Sterne et al., 2019) to assess the risk of bias of the primary outcome. We will report the inter-rater agreement in terms of percentage agreement and kappa.

***4. Data to calculate effect sizes***

We will extract data to calculate effect sizes (the number of patients randomized to each arm, the number of patients assessed, the number of remitters, the scale used, the mean, standard deviation and the number assessed for continuous outcomes) When only change from baseline to endpoint is reported for continuous outcomes, we will use it instead of endpoint mean.(Costa et al., 2013)

**Primary outcome and secondary outcomes**

The primary outcome of interest in this study is treatment efficacy at four weeks post‐treatment or at its closest time point. Intention-to-treat analysis will be prioritized whenever available. We will use the number of participants randomized as the denominator for dichotomous outcomes. We will use odds ratio for dichotomous outcomes, mean difference for continuous outcomes expressed in minutes and percent.

1. Efficacy: remission defined as reaching a satisfactory state at endpoint measured by any validated self-reported scale (dichotomous)

Secondary outcomes are as follows;

2. Acceptability: dropouts for any reason (dichotomous)

3. Sleep diary measures (continuous)

3.1. Sleep efficiency (SE, %)

3.2. Total sleep time (TST, min)

3.3. Sleep onset latency (SOL, min)

3.4. Wake after sleep onset (WASO, min)

4. Efficacy at long-term follow-up. (dichotomous, longest follow-up between 3 to 12 months)

**Hierarchy of outcome measures**

For efficacy, we will prioritize the remission using the Insomnia Severity Index (7 or less points at endpoint) (Morin et al., 2011) and its imputed number. If it is not reported, we will use the following scales in this order: the remission using the Functional Outcomes of Sleep Questionnaire-10 (18 or more points at endpoint) (Weaver et al., 2007), and then its imputed number; remission using the Epworth Sleepiness Scale (10 or less points at endpoint) (Johns, 1990), and then its imputed number; remission using the Pittsburgh Sleep Quality Index (5 or less points at endpoint) (Cole et al., 2006), and then its imputed number; remission using the Athens Insomnia Scale (5 or less points at endpoint) (Okajima et al., 2020), and then its imputed number; remission using any other validated self-reported scales; remission using sleep diary measures (both SOL and WASO less than 30 minutes at endpoint. When SOL and WASO are reported only separately, we will prioritize WASO. When only SOL is reported, we will use SOL.) (Edinger et al., 2009; Medicine, 2014) and its imputed number.

When any of the measure is reported using another definition of remission than stated above, we will use the definition stated by the authors. When any of the measure is reported only in continuous values, we will impute remission using mean and standard deviation. We will test the validity of this imputation method using studies that report the outcome both in continuous and dichotomous manner. When any of the measure is reported only in standardized mean difference, we will convert it into odds ratio using a validated method. (Chinn, 2000)

**Statistical analysis**

We will perform the analysis in *R* (latest version, R foundation, Vienna, Austria) (R Core Team, 2020) using *netmeta* (latest version) package (Rücker et al., 2020a) to conduct component network meta-analysis and *meta* (latest version) package (Balduzzi et al., 2019) to synthesize the outcomes in placebo arms and to assess the publication bias.

We will first perform a network meta-analysis lumping arms that include both cognitive and behavioral components as “cognitive-behavioral therapy,” those that involve cognitive but not behavioral components as “cognitive therapy” and those that involve behavioral but not cognitive components as “behavioral therapy” to gain a first insight of the relative treatment effects. (Table 2) We will examine the transitivity assumption by creating a table of important trial and patient characteristics to see if potential effect modifiers (publication year, proportion of patients with primary insomnia, age) are similarly distributed among comparisons. We will check the consistency of the network using local and global inconsistency tests.

Then we will perform component-level network meta-analysis. We will use a model that assumes additivity of components, i.e. assuming that the effect of combination therapy is the sum of the effects of its components. Given the expected clinical and methodological heterogeneity of treatment effects among the studies, we will use the random-effects model.

**Certainty of evidence**

We will assess the certainty of evidence in network estimates of the primary outcome using CINeMA. (Nikolakopoulou et al., 2020)

**Publication bias**

We will assess the presence of small study effects, including publication bias, in the evidence set by examining asymmetry in the contour-enhanced funnel plots of all active interventions (CBT, CT, BT) vs control condition (psychoeducation, waiting list, treatment as usual, attention or psychological placebo, no treatment) using the primary outcome.

**Sensitivity analyses**

1. Excluding studies without formal diagnosis of insomnia

2. Excluding studies focusing on patients with comorbidities (both physical and psychological)

3. Excluding studies with overall high dropout rate (20% or more)

4. Excluding studies at high overall risk of bias

5. Using completer-set analysis (dichotomous)

**Patient and public involvement**

There was no patient or public involvement in the development of this manuscript.

**Acknowledgements**

The views expressed are those of the authors and not necessarily those of affiliated organizations.

**Registration**

This protocol is prospectively registered in PROSPERO (CRD42022324233).

This research was prospectively registered (#2022033NIe), Ethical Committee, Faculty of Medicine, The University of Tokyo.

**Changes from the protocol (since the first registration on PROSPERO)**

25th May, 2022 (during screening, before data extraction). Although we stated in the protocol that "CBTI that clearly includes active components focusing on other symptoms, such as depression, anxiety or pain, will be excluded," in case the same intervention component was included in both arms, we decided to include the trial, as we will be able to see the additional benefit of CBTI.

**Support**

No financial support was used.

**Declarations of interest**

YF has received consultancy fee from Panasonic outside the submitted work.

MS reports personal fees from SONY outside the submitted work.

SF has a research grant from JSPS KAKENHI Grant Number JP 20K18964 and the KDDI Foundation.

SK has a research grant from Mental Health Okamoto Memorial Foundation and Fujiwara Memorial Foundation.

TAF reports grants and personal fees from Mitsubishi-Tanabe, personal fees from SONY, grants and personal fees from Shionogi, outside the submitted work; In addition, TAF has a patent 2020-548587 concerning smartphone CBT apps pending, and intellectual properties for Kokoro-app licensed to Mitsubishi-Tanabe.

EGO has received research and consultancy fees from Angelini Pharma. EGO is supported by the National Institute for Health Research (NIHR) Research Professorship to Professor Andrea Cipriani (grant RP-2017-08-ST2-006), by the National Institute for Health Research (NIHR) Applied Research Collaboration (ARC) Oxford and Thames Valley, by the National Institute for Health Research (NIHR) Oxford cognitive health Clinical Research Facility and by the NIHR Oxford Health Biomedical Research Centre (grant BRC-1215-20005)

OE was supported by the Swiss National Science Foundation (Ambizione grant number 180083)

MP reports no competing interest.

#### 1.2. Changes from the protocol.

25th May 2022 During screening, before data extraction. Although we stated in the protocol that "CBTI that clearly includes active components focusing on other symptoms, such as depression, anxiety or pain, will be excluded," in case the same intervention component was included in both arms, we decided to include the trial, as we would be able to see the additional benefit of CBTI.

4th August 2022. During screening, before data extraction, we decided to add “acceptance and commitment therapy” as eligible components. We decided to lump “acceptance and commitment therapy” together with the “mindfulness” component and call them “third-wave components”. We decided to exclude “imagery rehearsal” because it was primarily examined among PTSD patients and including this element would violate the transitivity assumption, which is critical for conducting network meta-analysis.

6th September 2022. During screening, before data extraction, we decided to use the Sleep Condition Indicator (remission: 17 or more points at endpoint) as an outcome measure and prioritise it next to ISI.

5th January 2023. During data extraction trial, we noticed that using the post-treatment time point allows for a clearer distinction from the long-term effect than using the 4-week post-treatment time point. We therefore decided to use the post-treatment time point for the primary outcome.

10th March 2023. We decided to use the baseline severity to test the transitivity assumption in addition to publication year, proportion of patients with primary insomnia and age.

18th March 2023. We decided not to conduct completer set analysis because such information was often not available.

19th April 2023. We decided to use 85% or more as the remission threshold for sleep efficiency.

23rd April 2023. We decided to conduct additional sensitivity analyses 1) focusing on trials using ISI as the outcome measure to see the influence of choice of measurements; 2) excluding delivery format components (ind, gp, ff, ae, he) to see the influence of extracting delivery methods as components; 3) treating insomnia severity as a continuous value instead of a binary outcome to examine the influence of dichotomizing the outcome; and 4) excluding arms with less than 10 participants to see the influence of small study effects.

#### 1.3. List of all variables for which data were sought

| **variable** | **description** |
| --- | --- |
| Year | Publication year of the primary report. If only the results from registry were available, we used the date when the results were posted. |
| Ind_clu | Unit of randomization. Individual or cluster. |
| ICC_for_cRCT | If it was a cluster RCT, we extracted Intra-cluster Correlation Coefficient. If it was not clearly stated, use “0.05”. |
| Country | Countries where the trial took place. |
| Single_multi | Whether the recruitment took place at a single or multi centers. |
| Age_mean | Mean age of the patients in the arm |
| Age_sd | Standard deviation of the age. |
| Age_n | The number of patients used for calculating Age_mean. |
| F_n | Number of females in the arm |
| Hyp_n | Number of patients using hypnotics. If the drug treatment was part of the protocol treatment, “Hyp_n” was equal to “n” (all the participants). If the drug treatment was not part of the protocol treatment but just allowed, we sued the number at baseline. We prioritised prescribed drugs to OTC drugs. |
| Insomnia diagnosis | If more than one formal diagnostic criteria were mentioned, prioritise DSM, ICSD and then ICD.  1. formal_DSM  1. formal_ICSD  1. formal_ICD  1. formal_Edinger2004  2. informal_ISI  2. informal_SCI  2. informal_PSQI  2. informal_AIS  2. informal_other_self_reported_scales  3. others |
| Primary insomnia_n | The number of primary insomnia (insomnia without comorbidities) patients in the arm. |
| Comorbidities | 0. primary_insomnia  1. primary_secondary_mixed  2. psy_MDD  2. psy_anxiety  2. psy_PTSD  2. psy_schizophrenia_or_psychosis  2. psy_bipolar  2. psy_substance_dependence  2. psy_others  2. psy_mixed  3. phy_dialysis  3. phy_heart_failure  3. phy_diabetes  3. phy_fibromyalgia  3. phy_cancer  3. phy_steroid  3. phy_pregnancy_perinatal  3. phy_menopause  3. phy_others  3. phy_mixed  4. psy_phy_mixed |

| **variable** | **description** |
| --- | --- |
| n | The number of patients **randomized** to the arm. |
| remission | The number of patients achieving remission at post-treatment or at its closest time point. If equidistant, we used the longer timeframe. (dichotomous) |
| def_remission_response | Definition of remission.. |
| scale used | The scale used for the definition of remission. Prioritize; ISI > SCI > FOSQ > ESS > PSQI > AIS > other_self_reported_scales > WASO_SOL > WASO > SOL. We will use the definition stated by the author. Note that we used sleep diary measures (subjective), not polysomnography measures (objective). |
| Severity_bl_mean | Mean severity measured at baseline. |
| Severity_bl_sd | Standard deviation of the above. |
| Severity_bl_n | The number of patients measured their severity at baseline. |
| Severity_ch_mean | Extracted only when Severity_ep_mean, Severity _ep_sd and Sevrity_ep_n were not available.  Mean change of Severity from baseline to the endpoint (post-treatment or at its closest time point). |
| Severity_ch_sd | Extracted only when Severity_ep_mean, Severity _ep_sd and Sevrity_ep_n were not available.  Standard deviation of the above. |
| Severity_ch_n | Extracted only when Severity_ep_mean, Severity _ep_sd and Sevrity_ep_n were not available.  The number of patients measured their severity change from baseline to the endpoint (post-treatment or at its closest time point). |
| Severity_ep_mean | Mean severity measured at post-treatment or at its closest time point. |
| Severity_ep_sd | Standard deviation of the above. |
| Severity_ep_n | The number of patients measured their severity at post-treatment or at its closest time point. |
| Severity_scale | The scale used for measuring the sleep-related severity. Prioritize; ISI > SCI> FOSQ > ESS > PSQI > AIS > other_self_reported_scales > WASO_SOL > WASO > SOL. We used the definition stated by the author. |
| dropout | The number of patients dropped out from the study for any reason (=the number of patients whose outcome data not available). Those who are lost to follow-up (assessment at post-treatment) were assumed to have dropped off. |
| SE_mean | Mean sleep efficiency in %. |
| SE_sd | Standard deviation of the above. |
| SE_n | The number of patients measured for the numbers above. |
| TST_mean | Mean total sleep time in minutes. |
| TST_sd | Standard deviation of the above. |
| TST_n | The number of patients measured for the numbers above. |
| SOL_mean | Mean sleep onset latency in minutes. |
| SOL_sd | Standard deviation of the above. |
| SOL_n | The number of patients measured for the numbers above. |
| WASO_mean | Mean wake after sleep onset in minutes. |
| WASO_sd | Standard deviation of the above. |
| WASO_n | The number of patients measured for the numbers above. |
| Sleep_diary_weeks | Timepoint when the sleep diary was measured |
| r_long | The number of patients achieving remission at long-term follow-up (longest follow-up between 3 to 12 months) |
| r_long_scale_used | Scales that were used for counting remission at long-term. |
| Severity_mean_long | Mean severity at long-term. |
| Severity_sd_long | Standard deviation of the above. |
| Severity_n_long | The number of patients measured for the numbers above. |
| Severity_ch_mean_long | Extracted this only when Severity_mean_long, Severity _ sd_long and Sevrity_n_long were not available. |
| Severity_ch_sd_long | Extracted this only when Severity_mean_long, Severity _ sd_long and Sevrity_n_long were not available. |
| Severity_ch_n_long | Extracted this only when Severity_mean_long, Severity _ sd_long and Sevrity_n_long were not available. |
| r_weeks_long | Timepoint of the measurement of the long-term follow-up in weeks. |

### **2. Screening process and results**

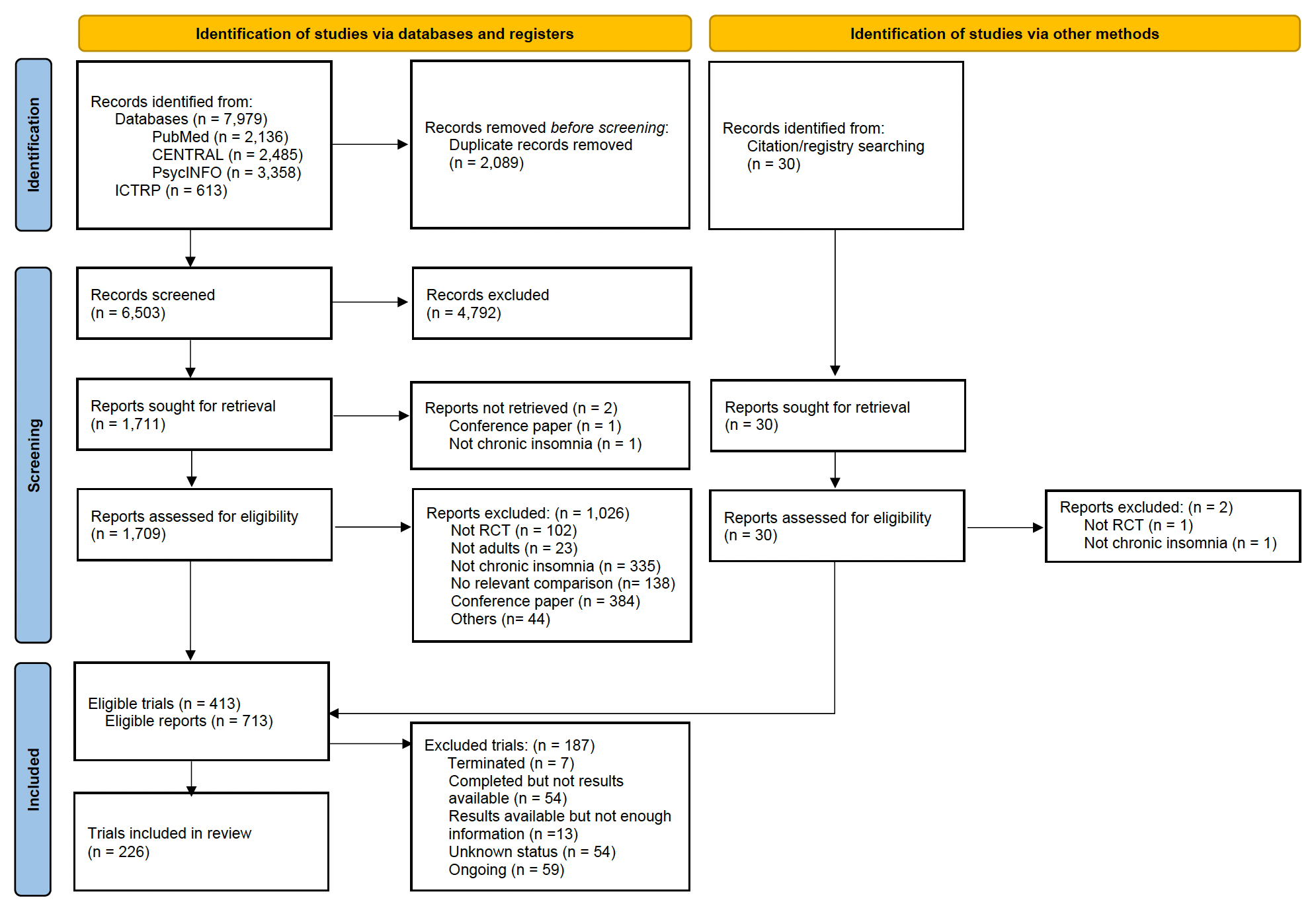

#### 2.1. Flow diagram

#### 2.2. List of the excluded studies with reasons (examples)

We excluded quasi-randomized trials, in which participants were not fully randomized or allocation was clearly not concealed.

- Sidani, S., Epstein, D. R., Fox, M., & Collins, L. Comparing the Effects of Single- and Multiple-Component Therapies for Insomnia on Sleep Outcomes. *Worldviews Evid.-Based Nurs*. 2019;16(3):195–203.
- Morgan, K., Dixon, S., Mathers, N., Thompson, J., & Tomeny, M. Psychological treatment for insomnia in the regulation of long-term hypnotic drug use. *Health technology assessment*. 2004;8(8):iii–68.
- Ellis JG, Cushing T, Germain A. Treating Acute Insomnia: A Randomized Controlled Trial of a "Single-Shot" of Cognitive Behavioral Therapy for Insomnia. *Sleep*. 2015;38(6):971-978.

We excluded trials when 1) the participants were clearly 17 years old or younger, or 2) some of the participants were 18 years old or older but the trial focused on adolescents. We included trials when some of the participants were 17 years old or younger but the majority of participants were 18 years old or older (mean – 2*SD >18).

- Hiscock H, Sciberras E, Mensah F, et al. Impact of a behavioural sleep intervention on symptoms and sleep in children with attention deficit hyperactivity disorder, and parental mental health: randomised controlled trial. *BMJ*. 2015;350:h68.
- Rogers VE, Zhu S, Ancoli-Israel S, Liu L, Mandrell BN, Hinds PS. A pilot randomized controlled trial to improve sleep and fatigue in children with central nervous system tumors hospitalized for high-dose chemotherapy. *Pediatr Blood Cancer*. 2019;66(8):e27814.

We included trials on patients with chronic insomnia disorder. We excluded trials when the eligibility criteria did not include significant distress or impairment in daytime functioning. (e.g. stating only “sleep problem” or “sleep disturbance”, even when they additionally used elevated scores on self-reported scales, or defining insomnia solely on sleep onset latency or wake time after sleep onset).

- Black, D. S., O'Reilly, G. A., Olmstead, R., Breen, E. C., & Irwin, M. R. Mindfulness meditation and improvement in sleep quality and daytime impairment among older adults with sleep disturbances: a randomized clinical trial. *JAMA internal medicine*. 2015;175(4):494–501.
- Malfliet, A., Bilterys, T., Van Looveren, E., Meeus, M., Danneels, L., Ickmans, K., Cagnie, B., Mairesse, O., Neu, D., Moens, M., Goubert, D., Kamper, S. J., & Nijs, J. The added value of cognitive behavioral therapy for insomnia to current best evidence physical therapy for chronic spinal pain: protocol of a randomized controlled clinical trial. *Brazilian journal of physical therapy*. 2019;23(1):62–70.

We included trials that used elevated scores on ISI for finding probable chronic insomnia disorder. Consequently, the minimum duration of the symptoms was 2 weeks and trials on patients suffering from sleep problems for less than 2 weeks were excluded.

- Wang LN, Tao H, Zhao Y, Zhou YQ, Jiang XR. Optimal timing for initiation of biofeedback-assisted relaxation training in hospitalized coronary heart disease patients with sleep disturbances. *J Cardiovasc Nurs*. 2014;29(4):367-376.

We were interested in dismantling the effects of each component of CBT-I and therefore excluded trials when they included active components focusing on other symptoms and those components were not equally distributed among arms.

- Thiart H, Lehr D, Ebert DD, Sieland B, Berking M, Riper H. Log in and breathe out: efficacy and cost-effectiveness of an online sleep training for teachers affected by work-related strain--study protocol for a randomized controlled trial. *Trials*. 2013;14:169.

We focused on the effects of CBT-I for improving insomnia symptoms, so we excluded trials that aimed at tapering hypnotics.

- Baillargeon L, Landreville P, Verreault R, Beauchemin JP, Grégoire JP, Morin CM. Discontinuation of benzodiazepines among older insomniac adults treated with cognitive-behavioural therapy combined with gradual tapering: a randomized trial. *CMAJ*. 2003;169(10):1015-1020.

### **3. Evaluation of components for each arm of included trials**

|  | **Percentage agreement** | **Kappa** |
| --- | --- | --- |
| **Educational components** |  |  |
| Sleep hygiene education (se) | 87.8% | 0.74 |
| Sleep diary (sd) | 80.3% | 0.43 |
| **Cognitive components** |  |  |
| Cognitive restructuring (cr) | 92.3% | 0.83 |
| Third wave components (th) | 96.5% | 0.76 |
| Constructive worry (cw) | 96.9% | 0.50 |
| **Behavioural components** |  |  |
| Sleep restriction (sr) | 92.7% | 0.85 |
| Stimulus control (sc) | 92.5% | 0.85 |
| Paradoxical intention (pi) | 98.8% | 0.79 |
| Relaxation (re) | 89.0% | 0.74 |
| **Others** |  |  |
| Non-specific treatment effect (ns) | 84.3% | 0.58 |
| Waiting component (w) | 94.4% | 0.81 |
| Conventional drug treatment (dt) | 73.1% | 0.45 |
| **Delivery methods** |  |  |
| Individual (ind) | 81.1% | 0.62 |
| Group (gp) | 93.7% | 0.77 |
| Face-to-face (ff) | 88.8% | 0.78 |
| Therapeutic guidance (tg) | 94.6% | 0.70 |
| Human encouragement (he) | 93.7% | 0.62 |
| Automated encouragement (ae) | 93.4% | 0.56 |

### **4. Examination of the imputation method**

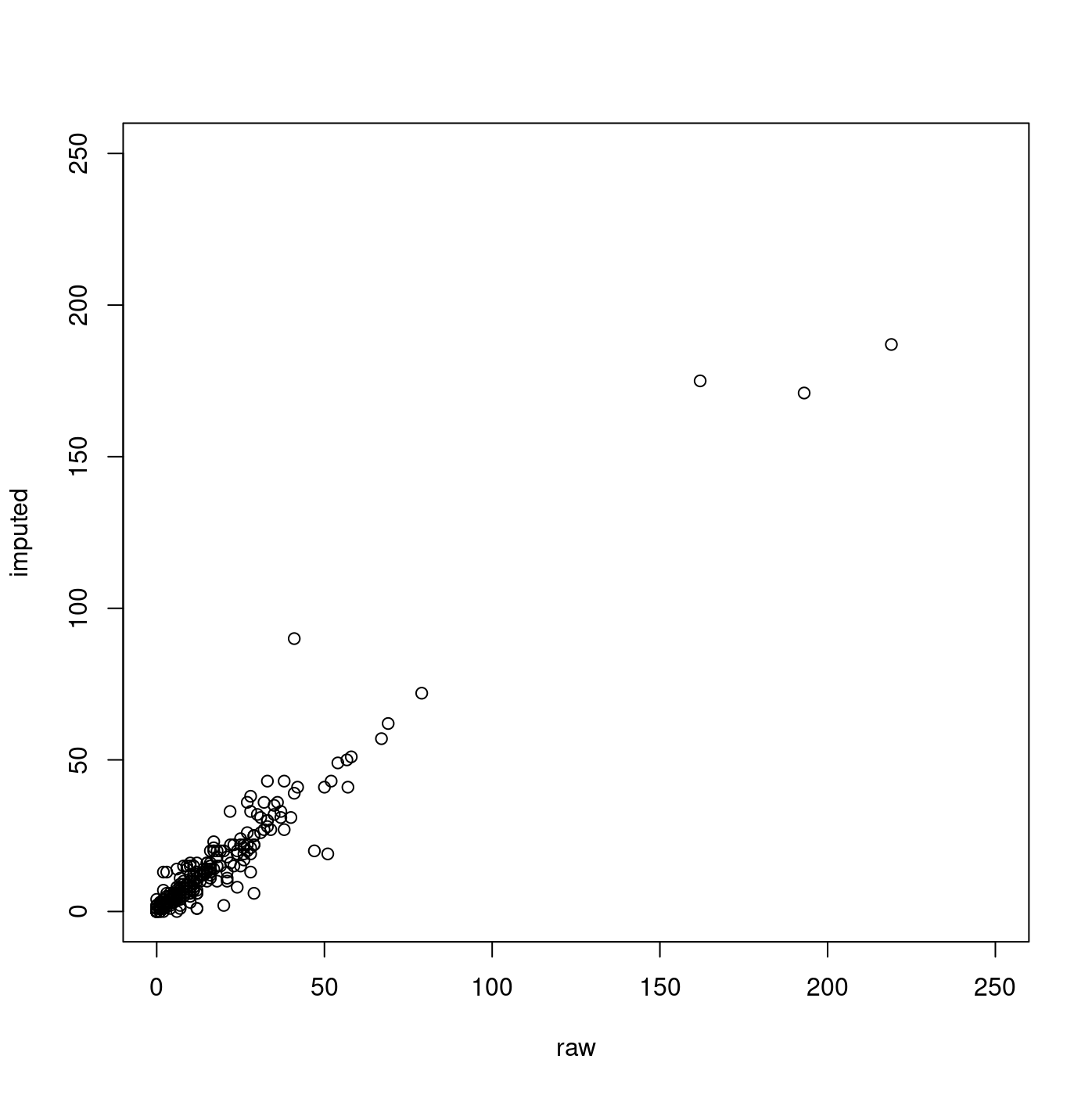

```
Single Score Intraclass Correlation

   Model: twoway
   Type : consistency

   Subjects = 267
     Raters = 2
   ICC(C,1) = 0.96

 F-Test, H0: r0 = 0 ; H1: r0 > 0
 F(266,266) = 49.5 , p = 6.15e-150

 95%-Confidence Interval for ICC Population Values:
  0.95 < ICC < 0.969
```

### **5. The revised Cochrane risk of bias**

We evaluated the risk of bias about the primary outcome, not the study quality, using the revised Cochrane risk-of-bias tool for randomized trials (RoB2). Here, we describe how we interpreted the signalling questions in each domain.

**Domain 1. Risk of bias arising from the randomization process**

1.1 Was the allocation sequence random?

1.2 Was the allocation sequence concealed until participants were enrolled and assigned to interventions?

We excluded studies where sequence generation was not clearly random, or where the allocation was clearly not concealed, were excluded.

1.3 Did baseline differences between intervention groups suggest a problem with the randomization process?

We evaluated if there were baseline differences in age, gender, and the primary outcome measure.

**Domain 2. Risk of bias due to deviations from intended interventions**

2.1. Were participants aware of their assigned intervention during the trial?

2.2. Were carers and people delivering the interventions aware of participants' assigned intervention during the trial?

In most cases, yes.

2.3. Were there deviations from the intended intervention that arose because of the trial context?

The term “trial context” refers to effects of recruitment and engagement activities on trial participants and when trial personnel undermine the implementation of the trial protocol in ways that would not happen outside the trial (e.g. daily practice). We expect non-adherence to the active treatment to occur outside the trials, too. We rated Yes only when deviations were more often in non-active arms, such as waiting list, treatment as usual or no treatment.

2.4 Were these deviations likely to have affected the outcome?

Yes.

2.5 Were these deviations from intended intervention balanced between groups?

If the proportions of deviations from intended intervention differed more than 10%, we rated No.

2.6 Was an appropriate analysis used to estimate the effect of assignment to intervention?

We used ITT or mITT in the primary analysis so we rated Yes.

**Domain 3. Risk of bias due to missing outcome data**

3.1 Were data for this outcome available for all, or nearly all, participants randomized?

In the guidance document of RoB2, it is stated that the proportion required for dichotomous outcomes depends on the risk of the event. As we expect the control event rate of 10-20% and the experimental event rate of 40-60%, we decided to use 10% as the threshold.

3.2 Is there evidence that the result was not biased by missing outcome data?

We rated yes when appropriate sensitivity analyses were conducted and they showed the result was not likely to be biased by missing outcome data.

3.3 Could missingness in the outcome depend on its true value?

Yes.

3.4 Is it likely that missingness in the outcome depended on its true value?

We rated yes when the proportions of missing data differed significantly (larger than 10%) between the arms or when the reasons for missing differed between the arms.

**Domain 4. Risk of bias in measurement of the outcome**

4.1 Was the method of measuring the outcome inappropriate?

Probably not.

4.2 Could measurement or ascertainment of the outcome have differed between intervention groups?

Probably not.

4.3 Were outcome assessors aware of the intervention received by study participants?

Yes.

4.4 Could assessment of the outcome have been influenced by knowledge of intervention received?

We rated no when more than two active comparators were used.

4.5 Is it likely that assessment of the outcome was influenced by knowledge of intervention received?

Probably not.

**Domain 5. Risk of bias in selection of the reported result**

5.1 Were the data that produced this result analyzed in accordance with a pre-specified analysis plan that was finalized before unblinded outcome data were available for analysis?

Is the numerical result being assessed likely to have been selected, on the basis of the results, from...

5.2. ... multiple eligible outcome measurements within the outcome domain?

5.3 ... multiple eligible analyses of the data?

We predefined in the protocol the hierarchy of outcome measures. We rated low risk of bias for the domain 5 when the trial reported the top priority outcome (ISI remission), or it reported ISI (continuous value) in a way that predefined in the statistical analysis plan. In most cases, details of statistical analysis plans were unavailable and we therefore rated some concerns.

**The inter-rater reliability of the assessment of risk of bias**

Overall risk of bias

|  | Low | Some | High |  | Percentage agreement |
| --- | --- | --- | --- | --- | --- |
| Low | 3 | 11 | 2 |  | 53.4% |
| Some | 16 | 59 | 32 |  | Weighed kappa |
| High | 2 | 25 | 39 |  | 0.33 |

Domain 1. Risk of bias arising from the randomization process

|  | Low | Some | High |  | Percentage agreement |
| --- | --- | --- | --- | --- | --- |
| Low | 72 | 12 | 3 |  | 74.1% |
| Some | 17 | 67 | 6 |  | Weighed kappa |
| High | 1 | 10 | 1 |  | 0.55 |

Domain 2. Risk of bias due to deviations from intended interventions

|  | Low | Some | High |  | Percentage agreement |
| --- | --- | --- | --- | --- | --- |
| Low | 143 | 18 | 9 |  | 77.8% |
| Some | 6 | 3 | 2 |  | Weighed kappa |
| High | 6 | 1 | 1 |  | 0.13 |

Domain 3. Risk of bias due to missing outcome data

|  | Low | Some | High |  | Percentage agreement |
| --- | --- | --- | --- | --- | --- |
| Low | 70 | 19 | 17 |  | 56.6% |
| Some | 15 | 14 | 5 |  | Weighed kappa |
| High | 10 | 16 | 23 |  | 0.39 |

Domain 4. Risk of bias in measurement of the outcome

|  | Low | Some | High |  | Percentage agreement |
| --- | --- | --- | --- | --- | --- |
| Low | 56 | 28 | 1 |  | 68.8% |
| Some | 26 | 73 | 1 |  | Weighed kappa |
| High | 2 | 1 | 1 |  | 0.36 |

Domain 5. Risk of bias in selection of the reported result

|  | Low | Some | High |  | Percentage agreement |
| --- | --- | --- | --- | --- | --- |
| Low | 59 | 30 | 1 |  | 63.0% |
| Some | 32 | 60 | 2 |  | Weighed kappa |
| High | 1 | 4 | 0 |  | 0.31 |

The overall agreement of the assessment was fair, which was not worse than the interrater reliability reported by systematic review experts.

- Minozzi S, Cinquini M, Gianola S, Gonzalez-Lorenzo M, Banzi R. The revised Cochrane risk of bias tool for randomized trials (RoB 2) showed low interrater reliability and challenges in its application. *J Clin Epidemiol*. 2020;126:37-44.

### **6. Treatment level network meta-analysis**

#### 6.1. Assessment of transitivity

Statistical heterogeneity of the treatment-level NMA was estimated to be 𝜏𝜏2=0.21. The global, design-by-treatment test for inconsistency gave Q=37.14, with 31 degrees of freedom, p-value=0.20. The local approach to inconsistency (back-calculation method) gave the following results:

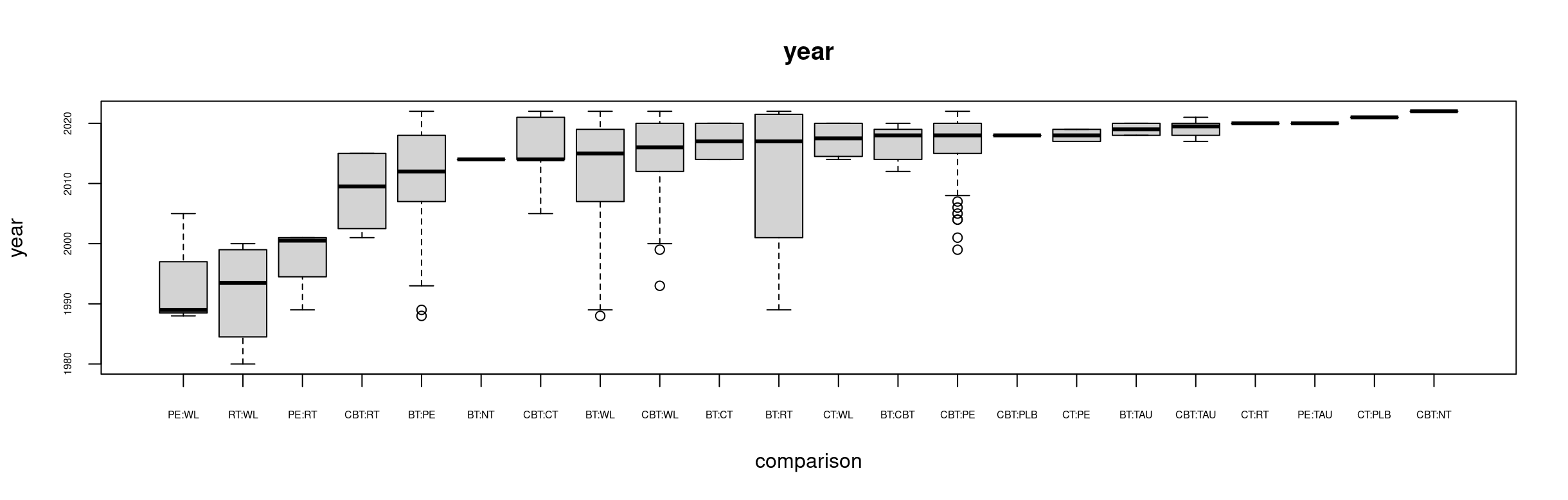

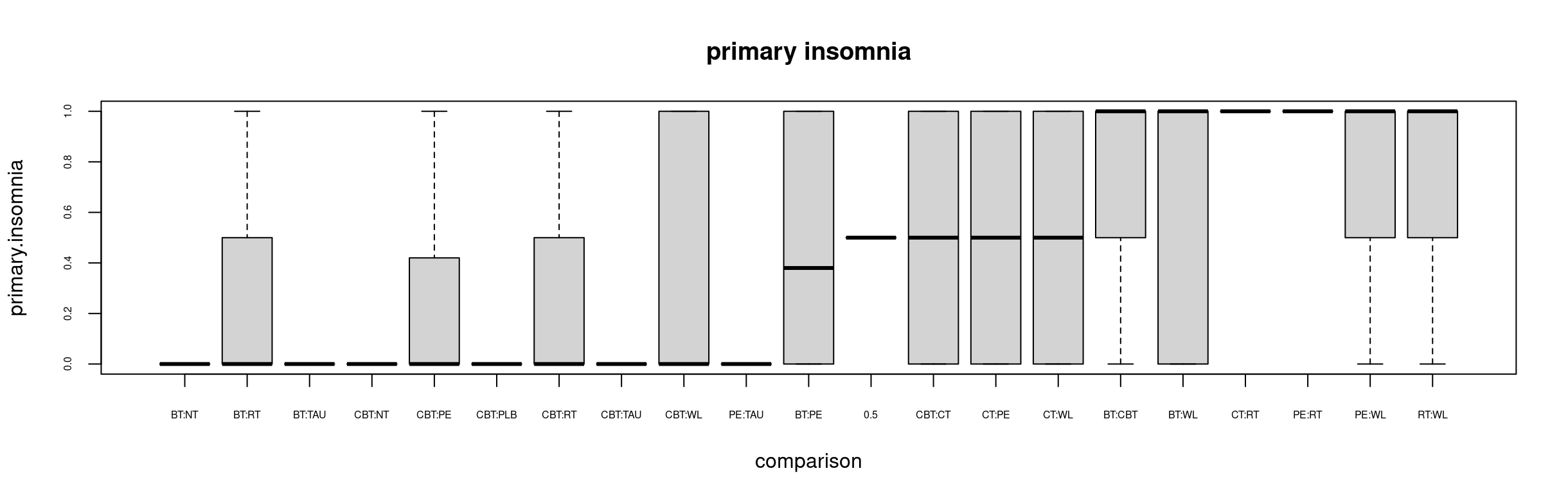

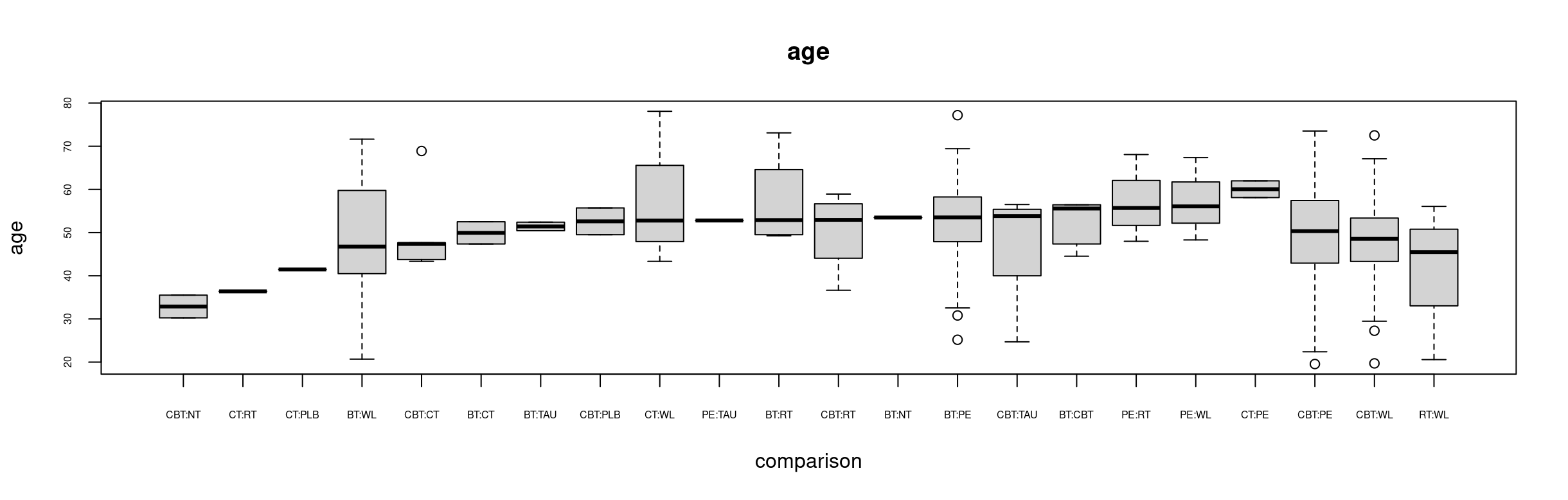

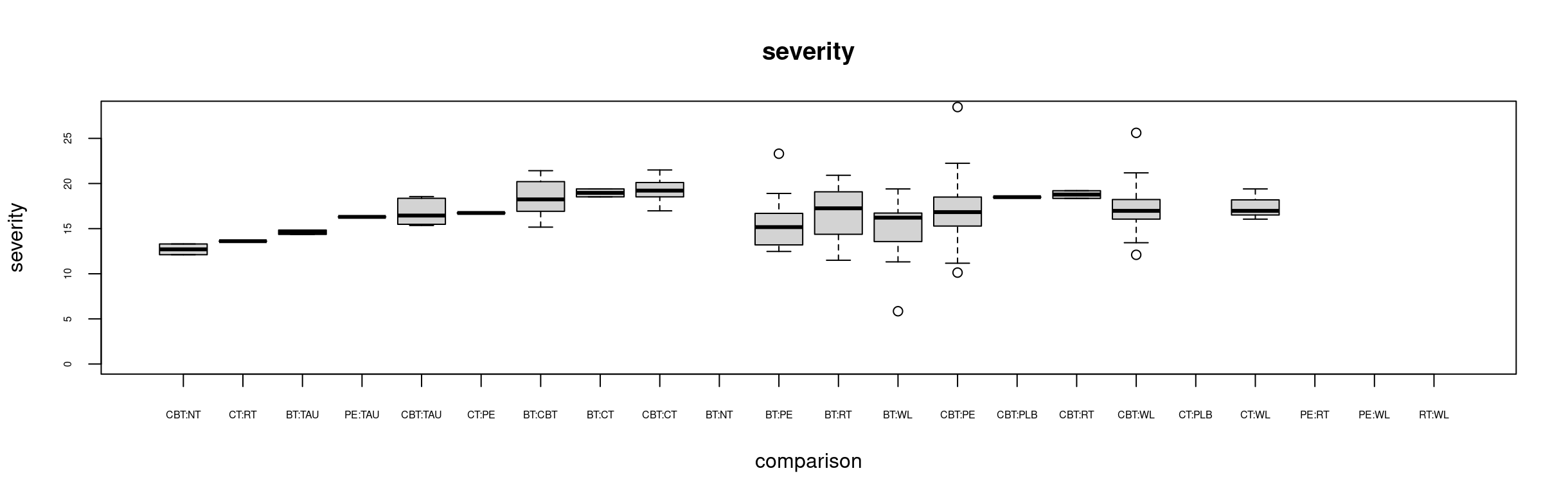
**6.2. Assessment of inconsistency for the primary outcome**

```
> # local
Separate indirect from direct evidence (SIDE) using back-calculation method

Random effects model:

 comparison  k prop     nma  direct  indir.    RoR     z p-value
     BT:CBT  5 0.18  0.7010  0.6871  0.7041 0.9759 -0.07  0.9421
      BT:CT  2 0.40  1.1535  1.3143  1.0565 1.2440  0.41  0.6838
      BT:NT  0    0  4.1134       .  4.1134      .     .       .
      BT:PE 25 0.46  2.5511  3.4841  1.9586 1.7789  2.17  0.0300
     BT:PLB  0    0 11.9325       . 11.9325      .     .       .
      BT:RT  4 0.34  1.9439  2.5177  1.6977 1.4830  0.84  0.4012
     BT:TAU  2 0.29  1.8485  1.5760  1.9731 0.7987 -0.39  0.6948
      BT:WL 22 0.52  3.7991  2.9919  4.9319 0.6066 -1.88  0.0595
     CBT:CT  5 0.43  1.6454  2.5128  1.1998 2.0943  1.44  0.1510
     CBT:NT  2 1.00  5.8675  5.8675       .      .     .       .
     CBT:PE 72 0.87  3.6390  3.4229  5.4176 0.6318 -1.85  0.0642
    CBT:PLB  2 0.56 17.0211 10.2840 32.1368 0.3200 -0.69  0.4916
     CBT:RT  4 0.36  2.7729  3.2544  2.5387 1.2819  0.55  0.5790
    CBT:TAU  6 0.75  2.6368  2.9361  1.9222 1.5275  0.76  0.4502
     CBT:WL 65 0.80  5.4193  5.7709  4.2008 1.3737  1.27  0.2053
      CT:NT  0    0  3.5661       .  3.5661      .     .       .
      CT:PE  2 0.17  2.2116  2.9536  2.0830 1.4180  0.51  0.6121
     CT:PLB  1 0.47 10.3448 19.0000  6.0801 3.1249  0.69  0.4916
      CT:RT  1 0.11  1.6853  1.2000  1.7567 0.6831 -0.38  0.7063
     CT:TAU  0    0  1.6026       .  1.6026      .     .       .
      CT:WL  4 0.21  3.2936  9.8876  2.4784 3.9895  2.13  0.0333
      NT:PE  0    0  0.6202       .  0.6202      .     .       .
     NT:PLB  0    0  2.9009       .  2.9009      .     .       .
      NT:RT  0    0  0.4726       .  0.4726      .     .       .
     NT:TAU  0    0  0.4494       .  0.4494      .     .       .
      NT:WL  0    0  0.9236       .  0.9236      .     .       .
     PLB:PE  0    0  0.2138       .  0.2138      .     .       .
      RT:PE  4 0.26  1.3123  1.6532  1.2107 1.3654  0.62  0.5337
     TAU:PE  1 0.05  1.3800  4.5000  1.2918 3.4836  1.10  0.2720
      WL:PE  3 0.03  0.6715  0.2318  0.6926 0.3347 -1.47  0.1415
     PLB:RT  0    0  0.1629       .  0.1629      .     .       .
    PLB:TAU  0    0  0.1549       .  0.1549      .     .       .
     PLB:WL  0    0  0.3184       .  0.3184      .     .       .
     RT:TAU  0    0  0.9509       .  0.9509      .     .       .
      RT:WL  4 0.21  1.9544  2.1505  1.9058 1.1284  0.22  0.8252
     TAU:WL  0    0  2.0552       .  2.0552      .     .       .

Legend:
 comparison - Treatment comparison
 k          - Number of studies providing direct evidence
 prop       - Direct evidence proportion
 nma        - Estimated treatment effect (OR) in network meta-analysis
 direct     - Estimated treatment effect (OR) derived from direct evidence
 indir.     - Estimated treatment effect (OR) derived from indirect evidence
 RoR        - Ratio of Ratios (direct versus indirect)
 z          - z-value of test for disagreement (direct versus indirect)
 p-value    - p-value of test for disagreement (direct versus indirect)
```

#### 6.3. League table for treatment-level NMA

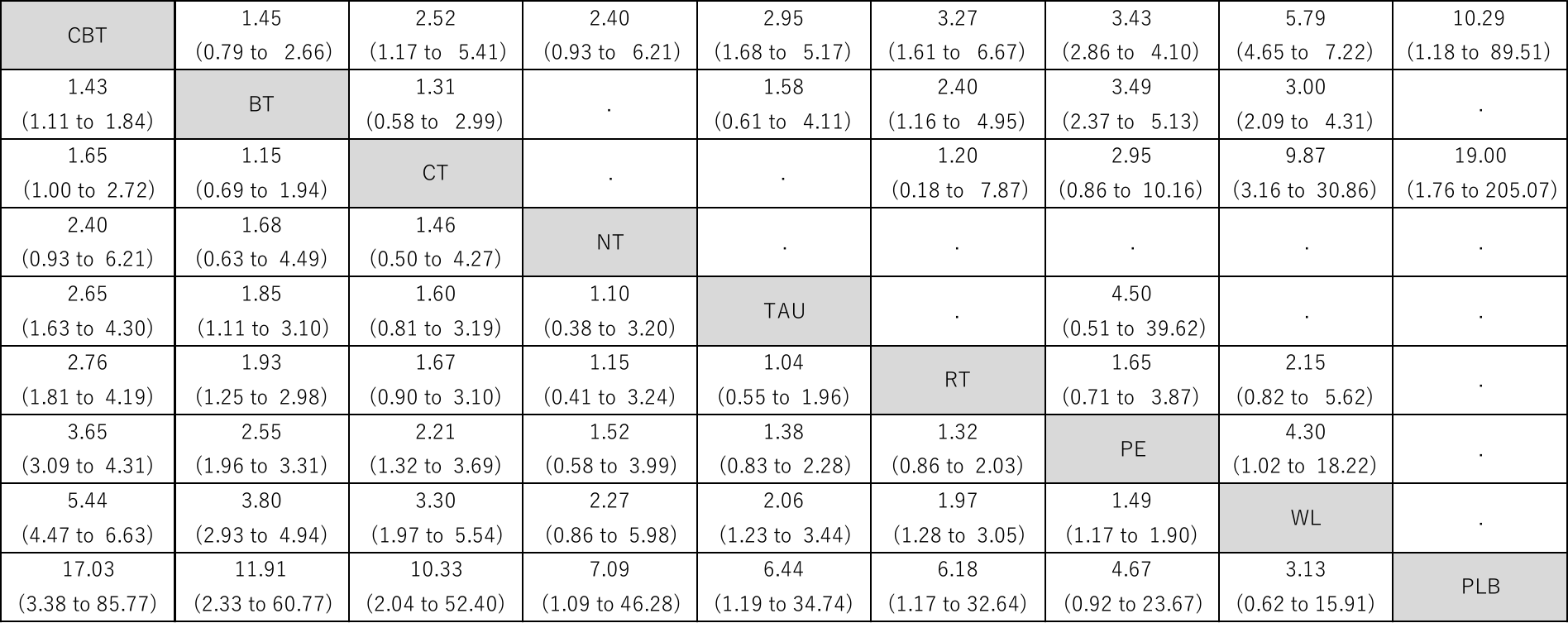

#### 6.4. Prediction intervals

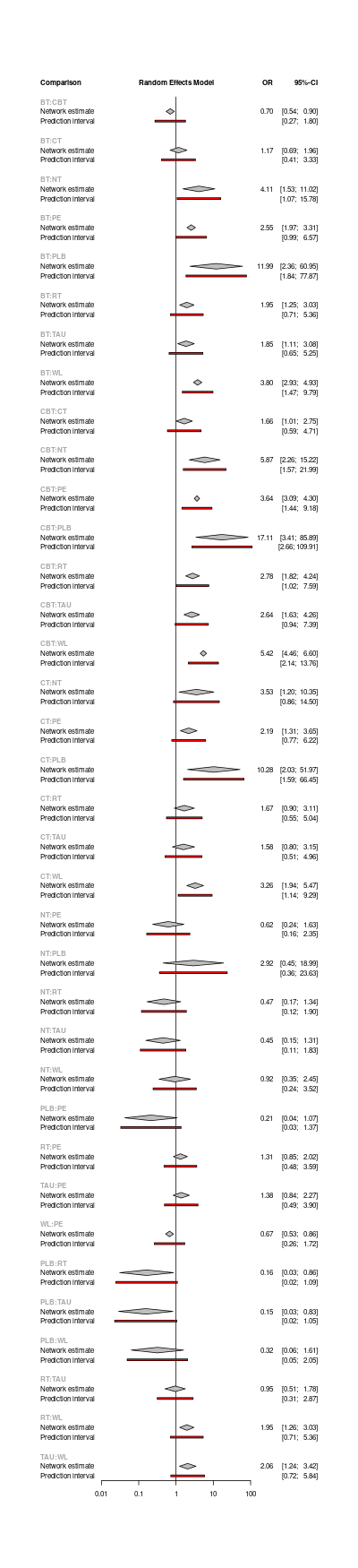

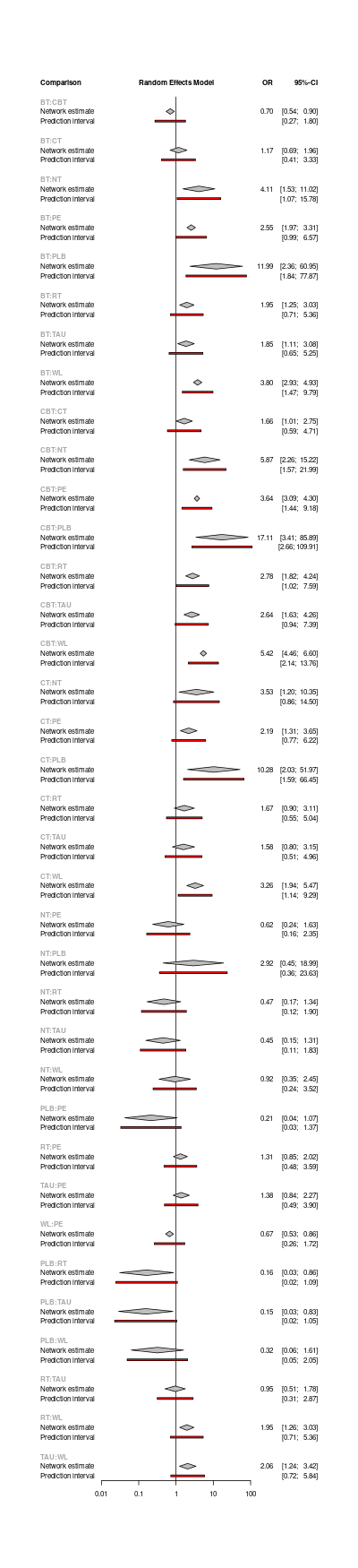

#### 6.5. Assessment of publication bias and small study effects

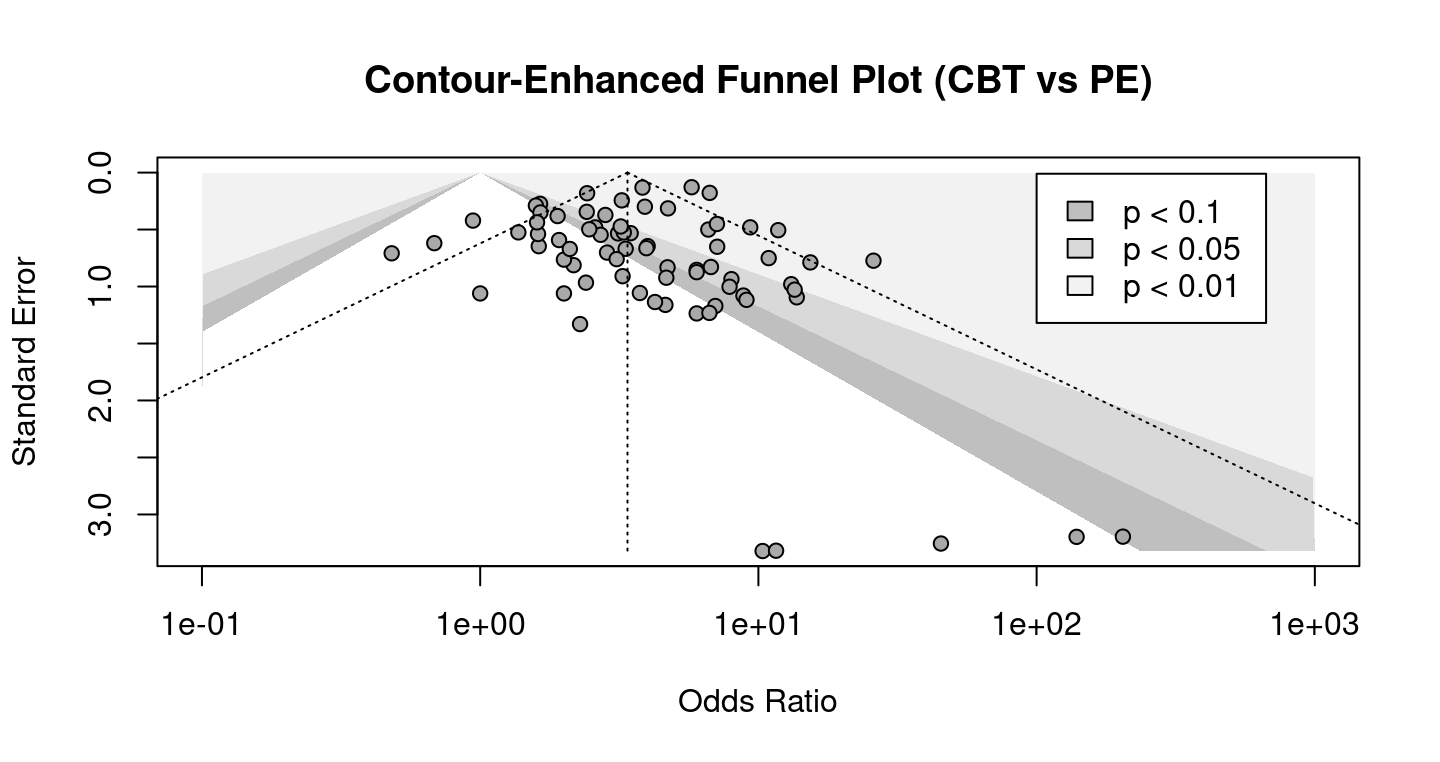

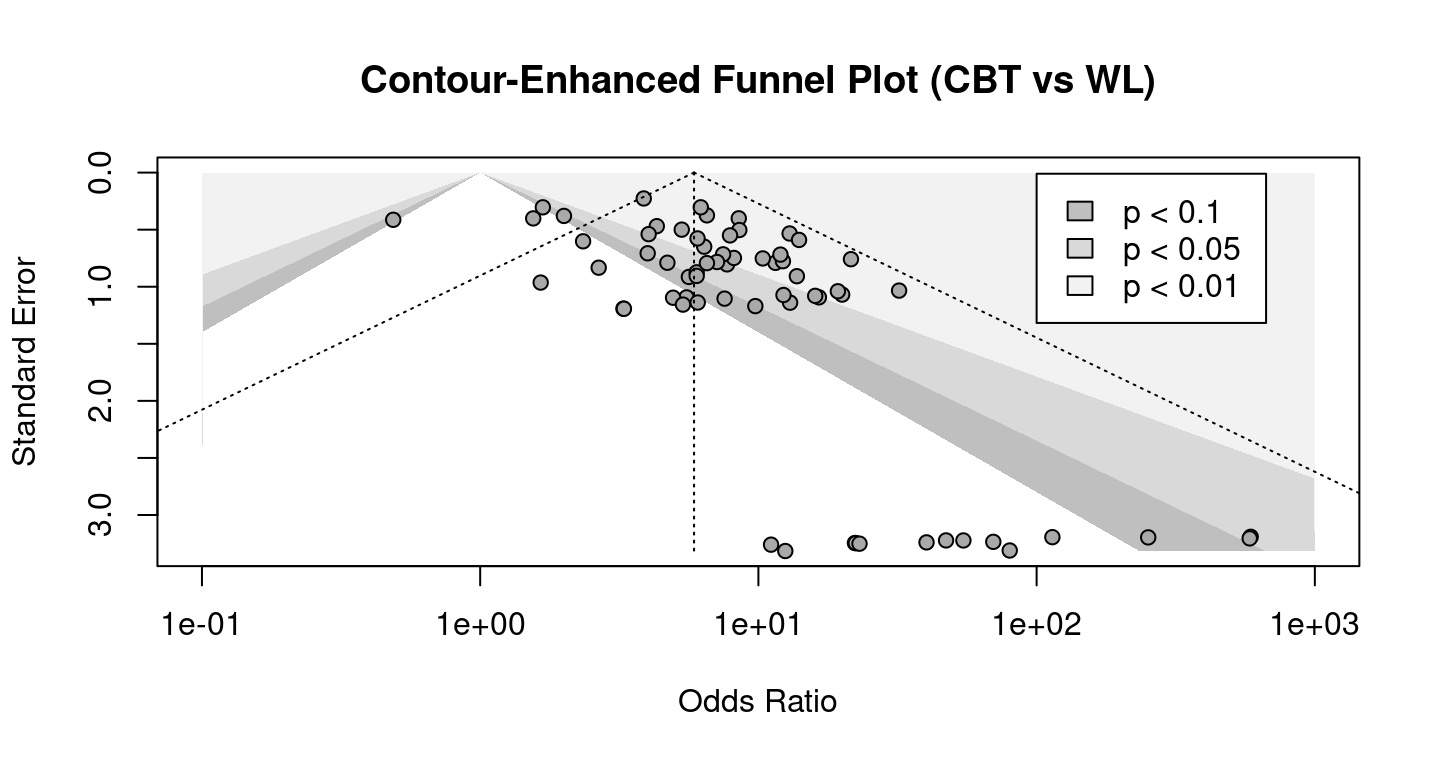

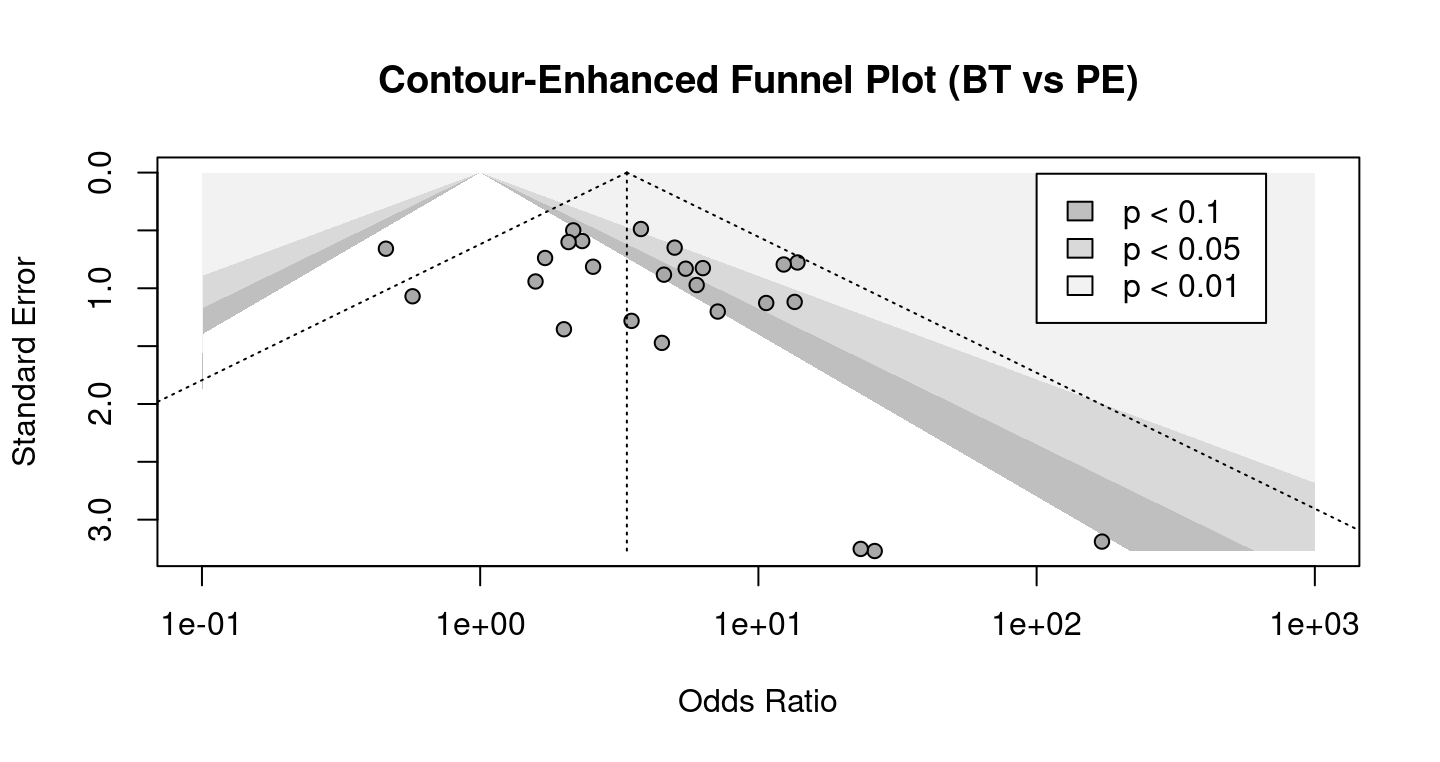

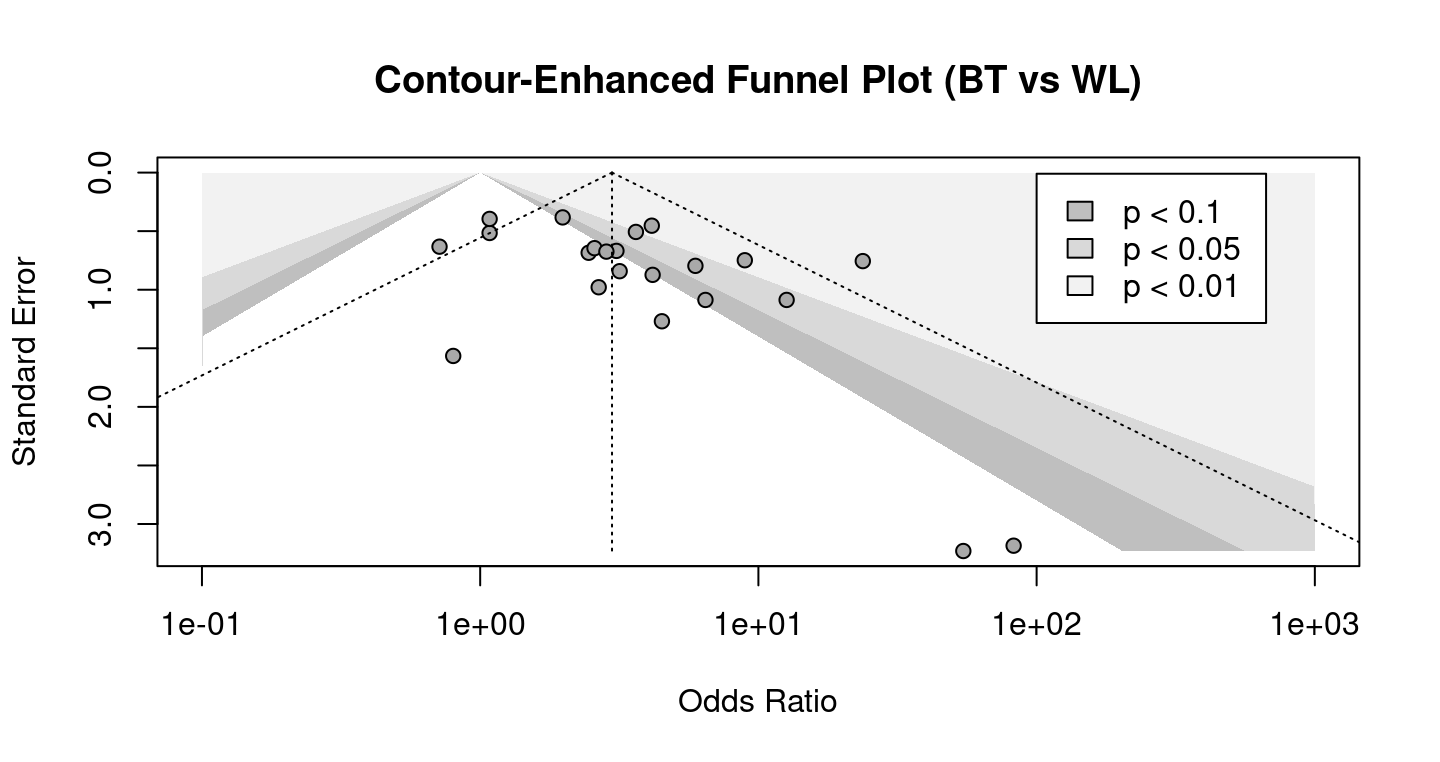

#### 6.6. CINeMA

Within-study bias: We used the majority RoB2 assessment.

Reporting bias: Low.

Indirectness: No concerns.

Imprecision, heterogeneity, incoherence: We used OR=1.5 as the clinically meaningful threshold.

Each domain: Major concerns=-3, some concerns=-1, low risk/no concerns=±0.

Confidence rating: 0=High, -1 to -3=moderate, -4 to -6=low, -7 or less=very low.

| **Comparison** | **k** | **Within-study bias** | **Reporting bias** | **Indirectness** | **Imprecision** | **Heterogeneity** | **Incoherence** | **Confidence rating** | **Reasons for downgrading** |
| --- | --- | --- | --- | --- | --- | --- | --- | --- | --- |
| BT:CBT | 6 | Some concerns | Low risk | No concerns | No concerns | Major concerns | No concerns | Low | ["Within-study bias","Heterogeneity"] |
| BT:CT | 2 | Some concerns | Low risk | No concerns | Some concerns | Some concerns | No concerns | Moderate | ["Within-study bias","Imprecision","Heterogeneity"] |
| BT:NT | 1 | Major concerns | Low risk | No concerns | No concerns | Some concerns | No concerns | Low | ["Within-study bias","Heterogeneity"] |
| BT:PE | 25 | Some concerns | Low risk | No concerns | No concerns | Some concerns | Some concerns | Moderate | ["Within-study bias","Heterogeneity","Incoherence"] |
| BT:RT | 4 | Some concerns | Low risk | No concerns | No concerns | Some concerns | No concerns | Moderate | ["Within-study bias","Heterogeneity"] |
| BT:TAU | 2 | Some concerns | Low risk | No concerns | No concerns | Major concerns | No concerns | Low | ["Within-study bias","Heterogeneity"] |
| BT:WL | 22 | Some concerns | Low risk | No concerns | No concerns | No concerns | No concerns | Moderate | ["Within-study bias"] |
| CBT:CT | 5 | Some concerns | Low risk | No concerns | No concerns | Major concerns | No concerns | Low | ["Within-study bias","Heterogeneity"] |
| CBT:NT | 2 | Major concerns | Low risk | No concerns | No concerns | No concerns | No concerns | Moderate | ["Within-study bias"] |
| CBT:PE | 72 | Some concerns | Low risk | No concerns | No concerns | No concerns | No concerns | Moderate | ["Within-study bias"] |
| CBT:PLB | 2 | Some concerns | Low risk | No concerns | No concerns | No concerns | No concerns | Moderate | ["Within-study bias"] |
| CBT:RT | 4 | Some concerns | Low risk | No concerns | No concerns | No concerns | No concerns | Moderate | ["Within-study bias"] |
| CBT:TAU | 6 | Some concerns | Low risk | No concerns | No concerns | Some concerns | No concerns | Moderate | ["Within-study bias","Heterogeneity"] |
| CBT:WL | 66 | Some concerns | Low risk | No concerns | No concerns | No concerns | No concerns | Moderate | ["Within-study bias"] |
| CT:PE | 2 | Some concerns | Low risk | No concerns | No concerns | Some concerns | No concerns | Moderate | ["Within-study bias","Heterogeneity"] |
| CT:PLB | 1 | Some concerns | Low risk | No concerns | No concerns | No concerns | No concerns | Moderate | ["Within-study bias"] |
| CT:RT | 1 | Some concerns | Low risk | No concerns | Some concerns | Some concerns | No concerns | Moderate | ["Within-study bias","Imprecision","Heterogeneity"] |
| CT:WL | 4 | Some concerns | Low risk | No concerns | No concerns | No concerns | Some concerns | Moderate | ["Within-study bias","Incoherence"] |
| PE:RT | 4 | Some concerns | Low risk | No concerns | Some concerns | Some concerns | No concerns | Moderate | ["Within-study bias","Imprecision","Heterogeneity"] |
| PE:TAU | 1 | Some concerns | Low risk | No concerns | Some concerns | Some concerns | No concerns | Moderate | ["Within-study bias","Imprecision","Heterogeneity"] |
| PE:WL | 3 | Some concerns | Low risk | No concerns | No concerns | Major concerns | No concerns | Low | ["Within-study bias","Heterogeneity"] |
| RT:WL | 4 | Some concerns | Low risk | No concerns | No concerns | Some concerns | No concerns | Moderate | ["Within-study bias","Heterogeneity"] |
| BT:PLB | 0 | Some concerns | Low risk | No concerns | No concerns | No concerns | Major concerns | Low | ["Within-study bias","Incoherence"] |
| CT:NT | 0 | Major concerns | Low risk | No concerns | Some concerns | No concerns | Major concerns | Very low | ["Within-study bias","Imprecision","Incoherence"] |
| CT:TAU | 0 | Some concerns | Low risk | No concerns | Some concerns | Some concerns | Major concerns | Low | ["Within-study bias","Imprecision","Heterogeneity","Incoherence"] |
| NT:PE | 0 | Major concerns | Low risk | No concerns | Major concerns | No concerns | Major concerns | Very low | ["Within-study bias","Imprecision","Incoherence"] |
| NT:PLB | 0 | Some concerns | Low risk | No concerns | Major concerns | No concerns | Major concerns | Very low | ["Within-study bias","Imprecision","Incoherence"] |
| NT:RT | 0 | Major concerns | Low risk | No concerns | Major concerns | No concerns | Major concerns | Very low | ["Within-study bias","Imprecision","Incoherence"] |
| NT:TAU | 0 | Major concerns | Low risk | No concerns | Major concerns | No concerns | Major concerns | Very low | ["Within-study bias","Imprecision","Incoherence"] |
| NT:WL | 0 | Major concerns | Low risk | No concerns | Major concerns | No concerns | Major concerns | Very low | ["Within-study bias","Imprecision","Incoherence"] |
| PE:PLB | 0 | Some concerns | Low risk | No concerns | Some concerns | No concerns | Major concerns | Low | ["Within-study bias","Imprecision","Incoherence"] |
| PLB:RT | 0 | Some concerns | Low risk | No concerns | No concerns | Some concerns | Major concerns | Low | ["Within-study bias","Heterogeneity","Incoherence"] |
| PLB:TAU | 0 | Some concerns | Low risk | No concerns | No concerns | Some concerns | Major concerns | Low | ["Within-study bias","Heterogeneity","Incoherence"] |
| PLB:WL | 0 | Some concerns | Low risk | No concerns | Major concerns | No concerns | Major concerns | Very low | ["Within-study bias","Imprecision","Incoherence"] |
| RT:TAU | 0 | Some concerns | Low risk | No concerns | Major concerns | No concerns | Major concerns | Very low | ["Within-study bias","Imprecision","Incoherence"] |
| TAU:WL | 0 | Some concerns | Low risk | No concerns | No concerns | Some concerns | Major concerns | Low | ["Within-study bias","Heterogeneity","Incoherence"] |

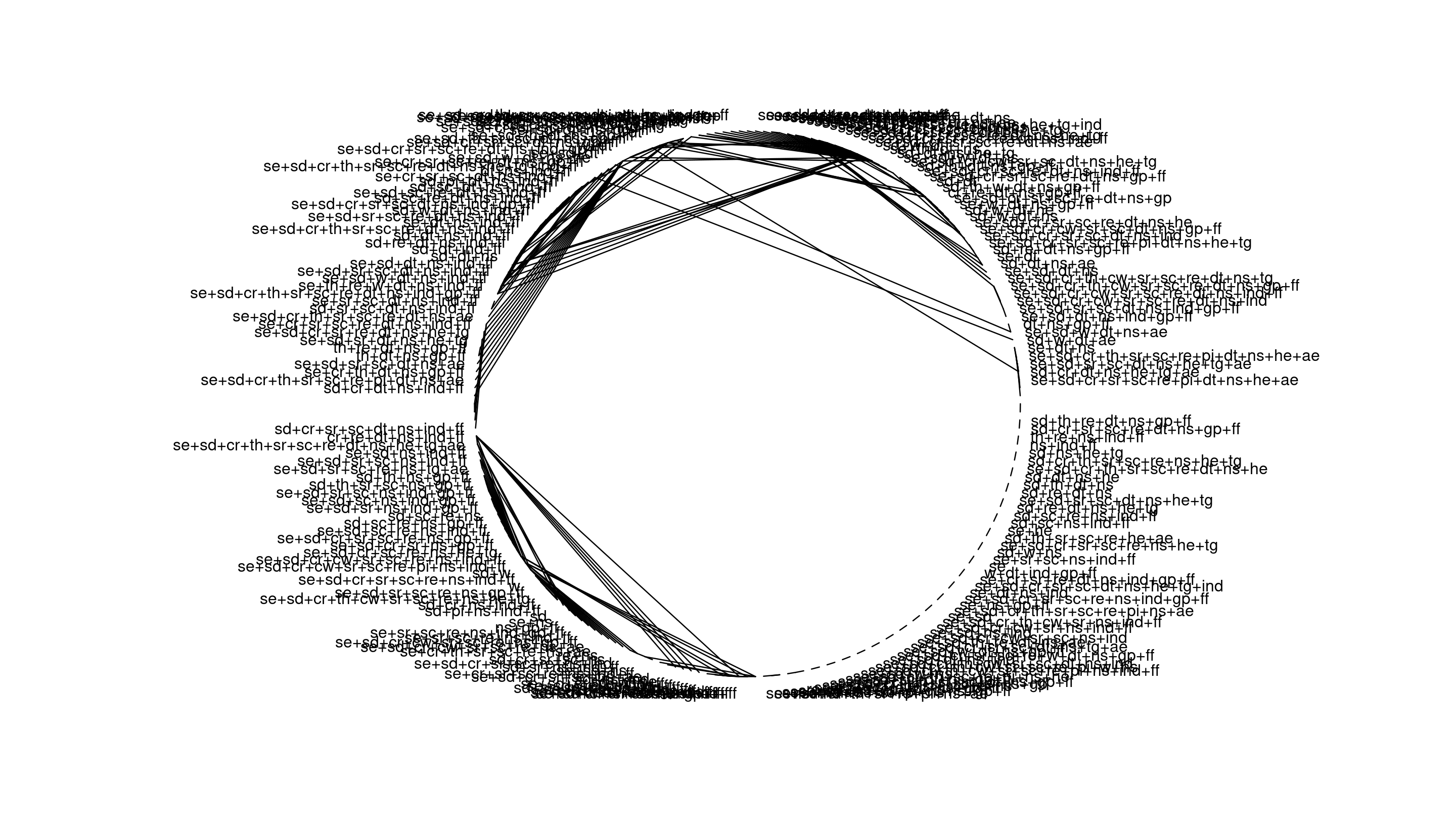

### **7. Component network meta-analysis**

#### 7.1. Network diagram

#### 7.2. The colouring scheme of the results

We used the procedure previously described in the so-called ‘Kilim plot’ to visualize the results.

• Seo M, Furukawa TA, Veroniki AA, et al. The Kilim plot: A tool for visualizing network meta-analysis results for multiple outcomes. *Res Synth Methods*. 2021;12(1):86-95. doi:10.1002/jrsm.1428

|  | Beneficial | Harmful |
| --- | --- | --- |
| p>0.10 |  |  |
| 0.05<p≤0.10 |  |  |
| 0.10<p≤0.05 |  |  |
| p≤0.01 |  |  |

#### 7.3. Sensitivity analyses

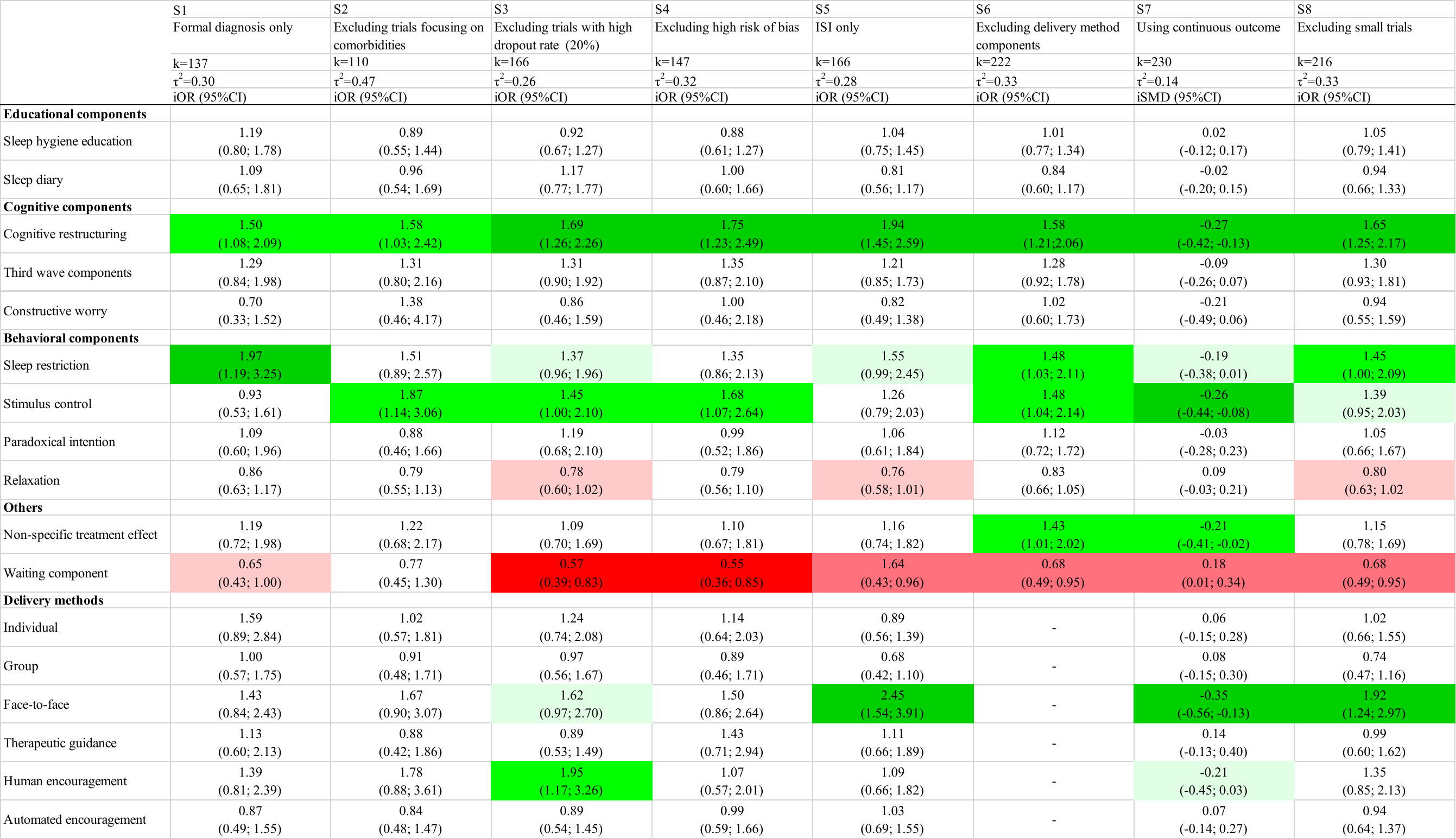

### **8. PRISMA-NMA**

| **Section/Topic** | **Item #** | **Checklist Item** | **Reported on Page #** |
| --- | --- | --- | --- |
| **TITLE** |  |  |  |
| Title | 1 | Identify the report as a systematic review *incorporating a network meta-analysis (or related form of meta-analysis).* | P1 |
| **ABSTRACT** |  |  |  |
| Structured summary | 2 | Provide a structured summary including, as applicable:  **Background:** main objectives  **Methods:** data sources; study eligibility criteria, participants, and interventions; study appraisal; and *synthesis methods, such as network meta-analysis.*  **Results:** number of studies and participants identified; summary estimates with corresponding confidence/credible intervals; *treatment rankings may also be discussed. Authors may choose to summarize pairwise comparisons against a chosen treatment included in their analyses for brevity.*  **Discussion/Conclusions:** limitations; conclusions and implications of findings.  **Other:** primary source of funding; systematic review registration number with registry name. | P4 |
| **INTRODUCTION** |  |  |  |
| Rationale | 3 | Describe the rationale for the review in the context of what is already known*, including mention of why a network meta-analysis has been conducted.* | P5 |
| Objectives | 4 | Provide an explicit statement of questions being addressed, with reference to participants, interventions, comparisons, outcomes, and study design (PICOS). | P5-6 |
| **METHODS** |  |  |  |
| Protocol and registration | 5 | Indicate whether a review protocol exists and if and where it can be accessed (e.g., Web address); and, if available, provide registration information, including registration number. | P5, appendix |
| Eligibility criteria | 6 | Specify study characteristics (e.g., PICOS, length of follow-up) and report characteristics (e.g., years considered, language, publication status) used as criteria for eligibility, giving rationale. *Clearly describe eligible treatments included in the treatment network, and note whether any have been clustered or merged into the same node (with justification).* | P5-6 |
| Information sources | 7 | Describe all information sources (e.g., databases with dates of coverage, contact with study authors to identify additional studies) in the search and date last searched. | P8 |
| Search | 8 | Present full electronic search strategy for at least one database, including any limits used, such that it could be repeated. | Appendix |
| Study selection | 9 | State the process for selecting studies (i.e., screening, eligibility, included in systematic review, and, if applicable, included in the meta-analysis). | P8, Appendix |
| Data collection process | 10 | Describe method of data extraction from reports (e.g., piloted forms, independently, in duplicate) and any processes for obtaining and confirming data from investigators. | P8 |
| Data items | 11 | List and define all variables for which data were sought (e.g., PICOS, funding sources) and any assumptions and simplifications made. | Appendix |
| **Geometry of the network** | **S1** | Describe methods used to explore the geometry of the treatment network under study and potential biases related to it. This should include how the evidence base has been graphically summarized for presentation, and what characteristics were compiled and used to describe the evidence base to readers. | Figure1,  Appendix |
| Risk of bias within individual studies | 12 | Describe methods used for assessing risk of bias of individual studies (including specification of whether this was done at the study or outcome level), and how this information is to be used in any data synthesis. | P8-9,  Appendix |
| Summary measures | 13 | State the principal summary measures (e.g., risk ratio, difference in means). *Also describe the use of additional summary measures assessed, such as treatment rankings and surface under the cumulative ranking curve (SUCRA) values, as well as modified approaches used to present summary findings from meta-analyses.* | P9 |
| Planned methods of analysis | 14 | Describe the methods of handling data and combining results of studies for each network meta-analysis. This should include, but not be limited to:   - *Handling of multi-arm trials;* - *Selection of variance structure;* - *Selection of prior distributions in Bayesian analyses; and* - *Assessment of model fit.* | P9 |
| **Assessment of Inconsistency** | **S2** | Describe the statistical methods used to evaluate the agreement of direct and indirect evidence in the treatment network(s) studied. Describe efforts taken to address its presence when found. | P9 |
| Risk of bias across studies | 15 | Specify any assessment of risk of bias that may affect the cumulative evidence (e.g., publication bias, selective reporting within studies). | P9 |
| Additional analyses | 16 | Describe methods of additional analyses if done, indicating which were pre-specified. This may include, but not be limited to, the following:   - Sensitivity or subgroup analyses; - Meta-regression analyses; - *Alternative formulations of the treatment network; and* - *Use of alternative prior distributions for Bayesian analyses (if applicable).* | P10 |
| **RESULTS†** |  |  |  |
| Study selection | 17 | Give numbers of studies screened, assessed for eligibility, and included in the review, with reasons for exclusions at each stage, ideally with a flow diagram. | Appendix |
| **Presentation of network structure** | **S3** | Provide a network graph of the included studies to enable visualization of the geometry of the treatment network. | Figure1 |
| **Summary of network geometry** | **S4** | Provide a brief overview of characteristics of the treatment network. This may include commentary on the abundance of trials and randomized patients for the different interventions and pairwise comparisons in the network, gaps of evidence in the treatment network, and potential biases reflected by the network structure. | P13, Figure1 |
| Study characteristics | 18 | For each study, present characteristics for which data were extracted (e.g., study size, PICOS, follow-up period) and provide the citations. | Appendix |
| Risk of bias within studies | 19 | Present data on risk of bias of each study and, if available, any outcome level assessment. | Appendix |
| Results of individual studies | 20 | For all outcomes considered (benefits or harms), present, for each study: 1) simple summary data for each intervention group, and 2) effect estimates and confidence intervals. *Modified approaches may be needed to deal with information from larger networks.* | ***Appendix*** |
| Synthesis of results | 21 | Present results of each meta-analysis done, including confidence/credible intervals. *In larger networks, authors may focus on comparisons versus a particular comparator (e.g. placebo or standard care), with full findings presented in an appendix. League tables and forest plots may be considered to summarize pairwise comparisons.* If additional summary measures were explored (such as treatment rankings), these should also be presented. | Figure2, Figure3, Appendix |
| **Exploration for inconsistency** | **S5** | Describe results from investigations of inconsistency. This may include such information as measures of model fit to compare consistency and inconsistency models, *P* values from statistical tests, or summary of inconsistency estimates from different parts of the treatment network. | Appendix |
| Risk of bias across studies | 22 | Present results of any assessment of risk of bias across studies for the evidence base being studied. | Appendix CINeMA |
| Results of additional analyses | 23 | Give results of additional analyses, if done (e.g., sensitivity or subgroup analyses, meta-regression analyses*, alternative network geometries studied, alternative choice of prior distributions for Bayesian analyses,* and so forth). | Appendix |
| **DISCUSSION** |  |  |  |
| Summary of evidence | 24 | Summarize the main findings, including the strength of evidence for each main outcome; consider their relevance to key groups (e.g., healthcare providers, users, and policy-makers). | P18 |
| Limitations | 25 | Discuss limitations at study and outcome level (e.g., risk of bias), and at review level (e.g., incomplete retrieval of identified research, reporting bias). *Comment on the validity of the assumptions, such as transitivity and consistency. Comment on any concerns regarding network geometry (e.g., avoidance of certain comparisons).* | P18 |
| Conclusions | 26 | Provide a general interpretation of the results in the context of other evidence, and implications for future research. | P19 |
| **FUNDING** |  |  |  |
| Funding | 27 | Describe sources of funding for the systematic review and other support (e.g., supply of data); role of funders for the systematic review. This should also include information regarding whether funding has been received from manufacturers of treatments in the network and/or whether some of the authors are content experts with professional conflicts of interest that could affect use of treatments in the network. | ***Y*** |
